# Estimating the global reduction in transmission and rise in detection capacity of the novel coronavirus SARS-CoV-2 in early 2020

**DOI:** 10.1101/2020.09.10.20192120

**Authors:** Antoine Belloir, François Blanquart

## Abstract

To better control the SARS-CoV-2 pandemic, it is essential to quantify the impact of control measures and the fraction of infected individuals that are detected. To this end we developed a deterministic transmission model based on the renewal equation and fitted the model to daily case and death data in the first few months of 2020 in 79 countries and states, representing 4.2 billions individuals. Based on a region-specific infection fatality ratio, we inferred the time-varying probability of case detection and the time-varying decline in transmissiblity. As a validation, the predicted total number of infected was close to that found in serosurveys; more importantly, the inferred probability of detection strongly correlated with the number of daily tests per inhabitant, with 50% detection achieved with 0.003 daily tests per inhabitants. Most of the decline in transmission was explained by the reductions in transmissibility (social distancing), which avoided 10 millions deaths in the regions studied over the first four months of 2020. In contrast, symptom-based testing and isolation of positive cases was not an efficient way to control the spread of the disease, as a large part of transmission happens before symptoms and only a small fraction of infected individuals was typically detected. The latter is explained by the limited number of tests available, and the fact that increasing test capacity increases the probability of detection less than proportionally. Together these results suggest that little control can be achieved by symptom-based testing and isolation alone.

## Introduction

The coronavirus SARS-CoV-2 originated in November-December 2019 (1), appeared as a cluster of cases of pneumonia of unknown etiology in the Wuhan province in China in December 2019-January 2020, and subsequently spread in the world in 2020. The rapid doubling time associated with the basic reproductive number R_0_ at 2-3 (2-4), together with the fact that an estimated ~50% of transmission is presymptomatic (5,6) make it difficult to control. A substantial proportion of infected individuals need to be hospitalised: 1 to 18% with increasing age in China, 4% overall in France (7-9). The infection fatality ratio (IFR) is around 1%, and much higher in the elderly (7-10).

By early March 2020, many regions of the world had imposed strong social distancing measures to reduce transmission and contain the spread of SARS-CoV-2. These social distancing measures were varied and included school closure, business closure, partial or full lockdowns, stay-at-home order, the prohibition of gatherings, curfews, etc. These measures resulted in the stabilisation or the inversion of the epidemic curve in many countries (11). This was accompanied by an increase in the capacity to PCR-test potentially infected individuals.

To improve the control of the epidemic, it is necessary to understand the transmission dynamics during the period of unrestricted growth in the first few months of 2020, and the impact of the subsequent reduction in transmission owing to (i) the depletion of susceptible individuals, (ii) the social distancing measures implemented, (iii) tests and isolation of cases. We develop a dynamical epidemiological model that describes the transmission dynamics with a discrete-time renewal equation. Thanks to published estimates of the IFR, our model predicts the daily number of all cases and the fraction of detected cases, and the daily number of deaths over the course of the epidemic and can thus be readily fit to data from 79 countries, states and provinces. Within each of these regions, we infer the time-varying probability of detection; the time-varying transmissibility; and we deduce the impact of detection and case isolation on transmission dynamics. The model is validated by the small difference between the predicted attack rate and that found in serological surveys. Finally, we show that the capacity to detect SARS-CoV-2 infections is strongly related to the number of tests performed per inhabitant, develop a novel model that relate the number of tests to the probability of detection and verify the model predictions. These results will serve to better understand and control transmission dynamics.

## Results

We model the dynamics of SARS-CoV-2 transmission for 79 geographical zones (countries, USA states, Canadian provinces and the Hubei province in China; hereafter “regions”) with a discrete-time renewal equation that describes how individuals are infected each day by transmission from previously infected individuals (Methods). Our model is akin to an existing model that predicts the daily number of deaths (11). The adapted renewal equation we use predicts in a deterministic way the daily numbers of infected, cases recorded, and deaths, given temporal profiles of transmissibility and case detection.

Infected individuals may die with a constant probability called the infection fatality ratio (IFR). We fix both the IFR and the distribution of time to death to values previously estimated from data from mainland China (8). The inference of the number of infected and hence the probability of detection crucially relies on the IFR, which links the daily deaths with the past number of infected individuals. The IFR is difficult to estimate because case detection is biased towards more severe cases. Early estimates relied on settings where tests were exhaustive such as repatriation flights or the Diamond Princess cruise boat (7,8,12). We use one of the published estimates of age-dependent IFR ((12); similar to other estimates, Supplementary Fig. 1) to compute a region-specific IFR that takes into account the regional age distribution. This region-specific IFR ranges from 0.3-0.4% (Bangladesh, Egypt, Pakistan, Philippines, South Africa) to 1.2%-1.4% (Germany, Italy, Portugal, Spain), and is typically around 1% in the regions examined (median 0.94%). This region-specific IFR does not take into account diffeences in health care capacity that could introduce additional variability (13). The IFR and the distribution of the time from infection to death allows us to project back in time the number of infected individuals.

We fit jointly the number of cases and deaths. This strategy has two advantages. While the number of deaths may be small, the number of cases is typically much larger and less subject to stochastic fluctuations. Furthermore, cases give an early signal of potential changes in transmissibility, as infected individuals may be detected as soon as symptoms occur, about a week after infection, while death occurs about three to four weeks after infection on average. The number of recorded cases, however, depends on the intensity of testing and the testing strategy. We account for changes in intensity of testing by modelling sand inferring a time-varying probability of case detection. We can thus interpret the number of cases recorded jointly with the number of deaths. Case detection is assumed to happen a few days after symptom onset (2.2 days on average), as inferred from (14), and to be followed by perfect isolation. Isolation reduces the pool of infected individuals who contribute to transmission (Methods).

From the renewal equation framework predicting the daily number of cases and deaths, we infer the time-varying transmission rate and the time-varying detection probability in the 79 studied regions. The chosen regions are those where the daily death incidence had reached 10 deaths at least once as of 23^rd^ April 2020 according to the John Hopkins Coronavirus Resource Center database. We fit the model by maximum likelihood to the case and death count data assuming the data points each day are drawn in a negative binomial distribution with mean given by the model prediction, and with an inferred dispersion parameter.

We validate our projections by comparing the inferred total attack rates—the proportion of individuals in the population that have ever been infected at a given date—with the number of infected individuals in nine regions where the number of infected at a certain time is known by systematic survey on a representative sample. The attack rate is given by the result of seroprevalence surveys, where a seropositive individuals is assumed to have been infected no later than 13 days in the past, corresponding to the median time to seroconversion (15). Note that in one case (Austria) we use results from a systematic PCR test survey. In that one case a positive individual is assumed to have been infected in the interval from 20 to 4 days ago (16) The attack rate predicted by our analysis was generally close to that in the data (Fig. 1), with no systematic bias. Countries above the identity line have more positive individuals in reality than predicted by the model. For these countries the true IFR is lower than the one assumed: given the realised number of deaths, the country actually had more infected individuals than what the model predicts. On the contrary, countries below the identity line have a higher IFR than the one assumed. Deviations of the true IFR from that assumed in the model bias the estimated absolute value of the detection probability, but not the temporal trends in the detection probability.

**Figure 1:**
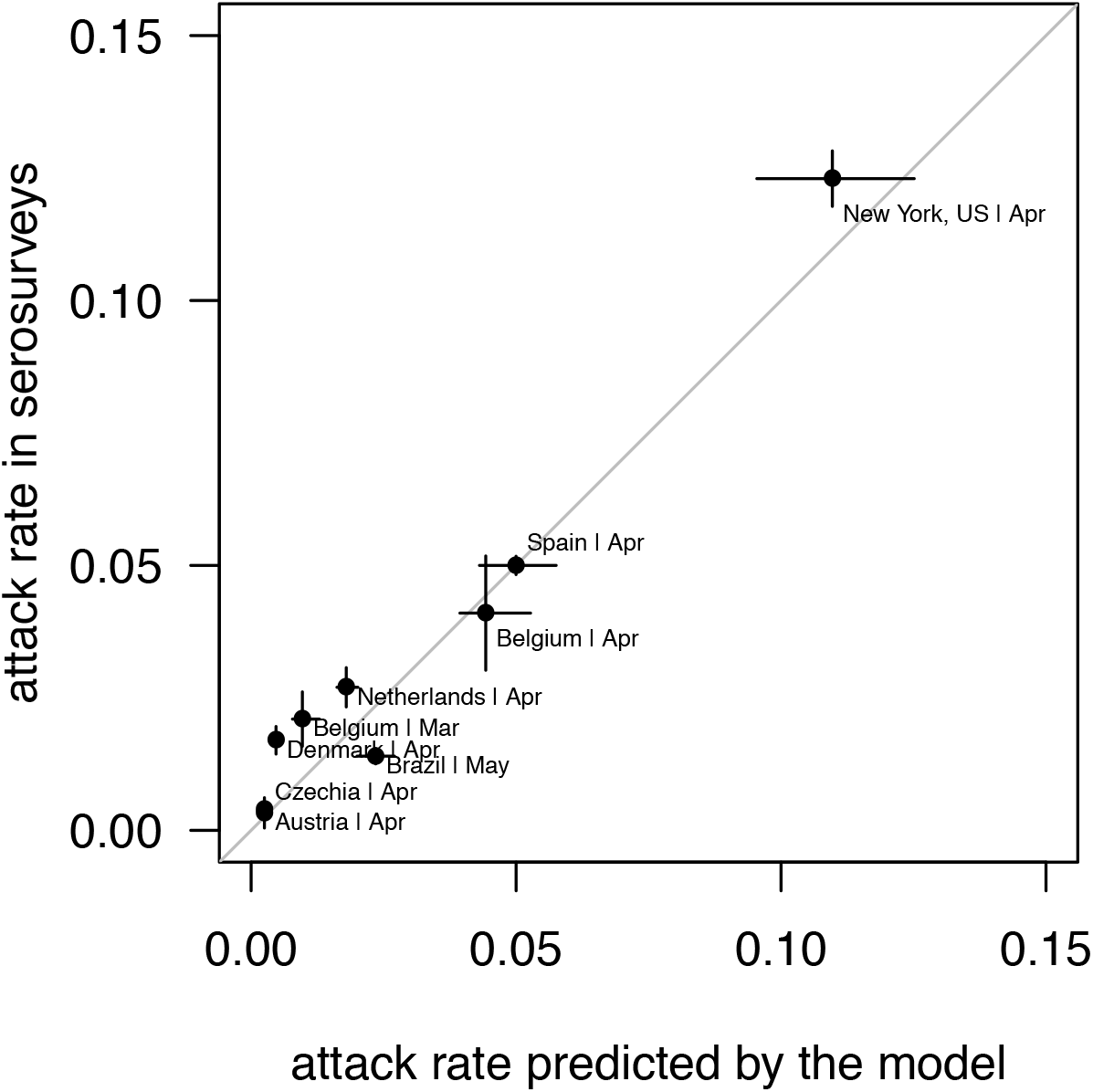
Comparison of the total number of infected (attack rate) found in systematic serological test surveys with that predicted by our model. The segments are 95% confidence intervals (for the data, binomial confidence intervals; for the model, estimated from the MCMC sample). Binomial confidence intervals for the data do not take into account the uncertainty on the representativity of the sample and could thus be underestimates. We used the model with the smooth sigmoid reduction in transmission; the model with the sharp transition gave very similar results.

To study the change in transmission following social distancing measures, one could infer the effects of different types of measures such as business, school, bar and restaurant closures, banning large gatherings, lockdowns, etc. However, these measures and their implementations are very varied across regions and multiple measures are often implemented simultaneously and may be accompanied by undocumented behavioural changes, complicating the inference of effects of individual measures (11). Instead, we estimate a region-specific reduction in transmissibility. We test two functional forms for the decline in transmissibility: (i) a sharp reduction in transmissibility at the date of social distancing, (ii) a smooth sigmoid reduction in transmissibility. For the first functional form, we considered the date of the national lockdownn (65/79 regions). If there was no national lockdown, we chose the date of regional lockdowns (5 regions: Algeria, Brazil, Indonesia, Oklahoma, Russia) or the date of a variety of distancing measures without strict lockdown (9 regions: British Columbia, Canada, Chile, Dominican Republic, Egypt, Iran, Ontario, Sweden, Turkey). When comparing the fit of the two functional forms with the Akaike Information Criterion (AIC), the smooth reduction in transmissibility fitted data better (an AIC difference greater than 4) in 50 regions out of 79. In these cases the reduction in transmissibility predated the date of social distancing by 5 to 20 days (Supplementary Fig. 3). In the 29 other regions, both functional forms were similar (Supplementary Fig. 2).

In most regions, we find a strong reduction in transmissibility accompanied by an increase in detection capacity. The basic reproduction number decreased from 3.7 on average across countries at the first date when 5 daily cases were reached, to 0.98 as of 8^th^ of May (Fig. 2B). There is substantial variation in the inferred initial transmissibility across regions. The mean probability of detection increased from 4% to 29% over the same period (Fig. 2C). The transmissibility remained above 1 (the threshold above which the epidemic expands in the absence of other measures) in several regions as of 8^th^ May, including Minnesotta, Brazil, Mexico, Pakistan, South Africa (Fig. 3A). The type of social distancing measure (national lockdown, regional lockdown, distancing) did not affect the final transmissibility (linear model for the final transmissibility as a function of the distancing measure; p= 0.46). The probability of detection as of 8^th^ May was below 50% for 67 out of 79 regions (Fig. 2B). The model predicted an attack rate of infection across regions of 0.1% (India) to 15% (New Jersey, USA).

**Figure 2:**
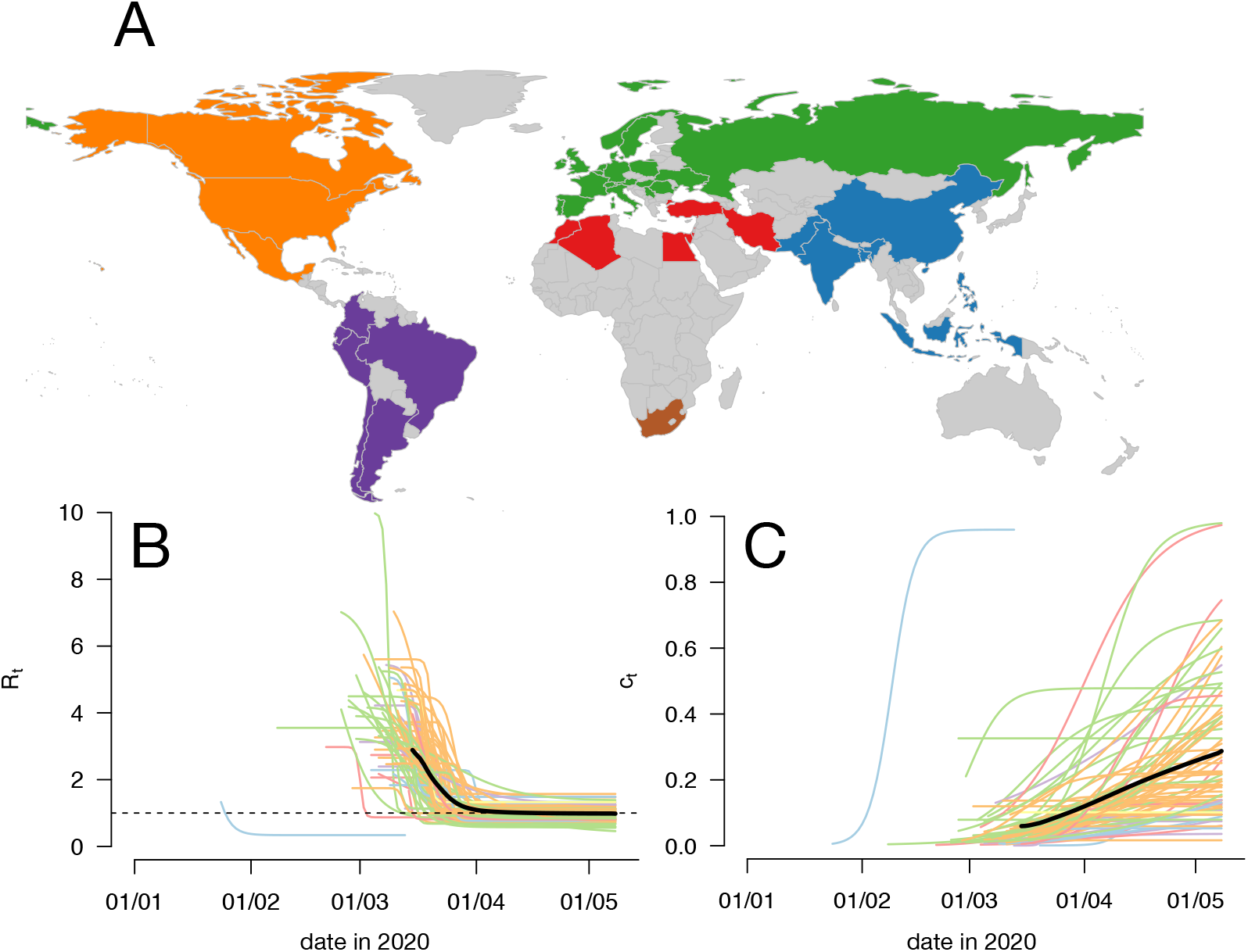
**Panel A** shows the map of the regions considered in this study, colored by geographic areas (Europe+Russia, North Africa/Middle East, Asia, South Africa, Central-South America, North America). The USA are represented by 33 states. China is represented by the Hubei province. **Panels B, C** show the inferred transmissibility and the probability of detection as a function of time for all regions. The overall mean is a thick black line. The early blue trajectory is that of the Hubei province in China.

**Figure 3:**
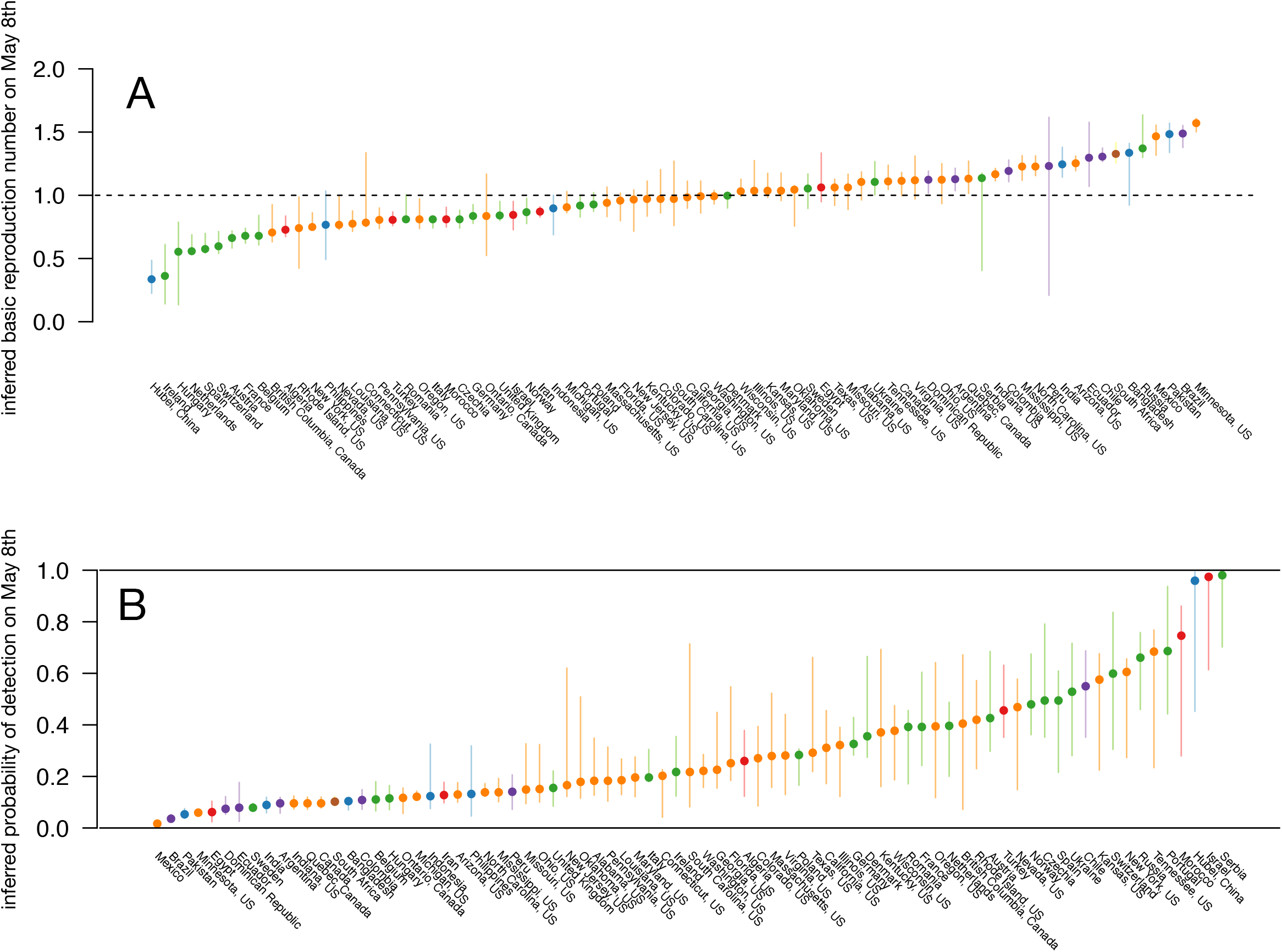
Inferred transmissibility R_t_ (**panel A**) and probability of detection C_t_ (**panel B**) as of May 8th, for each region. The point is the maximum likelihood estimate and the segment shows the 95% confidence intervals.

### Factors contributing to the reduction to transmission

The effective reproduction number 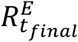 on the 8^th^ of May (*t_final_*), including the impacts of detection and isolation and immunity may be written as the product of the initial basic reproduction 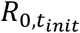 number times three factors that all reduce transmission:

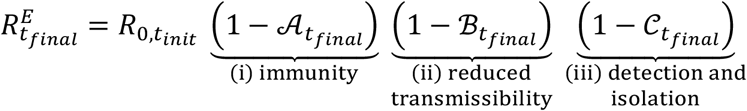

with 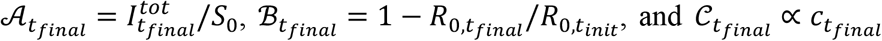 (Material and Methods). The reduction in overall transmission depends on (i) the depletion of the pool of susceptibles, (ii) reduced transmissibility impacting the basic reproduction number *R*_0_,*_t_*, (iii) testing and case isolation. We found that the factor contributing most to reduced transmission is the reduced transmissibility (Fig. 4B).

**Figure 4:**
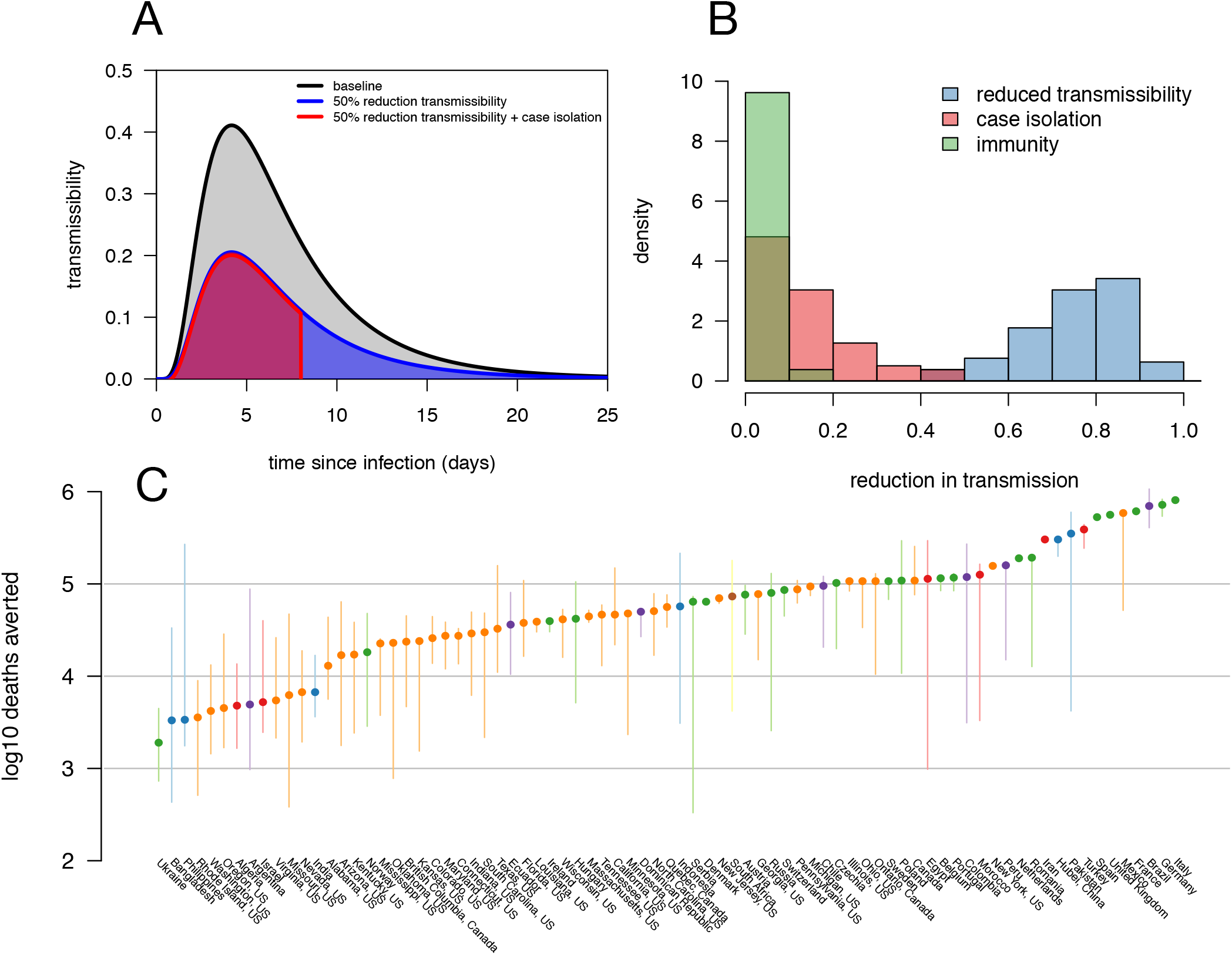
Impact of control measures and immunity on transmission dynamics. **Panel A** illustrates how social distancing and case isolation reduce transmission of the disease. The basic reproduction number is given by the area under the curve. A reduction in transmissibility uniformly reduces the R_t_ (blue curve and area), while detection and case isolation truncates the serial interval (red curve). **Panel B** represents the distribution of the reduction in transmission caused by social distancing (blue), detection and isolation (red) and immunity of already infected individuals (green) across the 79 regions. **Panel C** represents the logio number of deaths averted by social distancing between the beginning of the epidemic and the 8th May 2020.

The reduction in transmission owing to population immunity depends on the total number of individuals ever infected 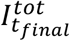 (the attack rate) over the initial number of susceptible individuals *S*_0_, assumed to be the total population size of the region. The attack rate was smaller than 2% in 47 regions out of 79. The reduction in the number of susceptible individuals that could lead to herd immunity is thus very small in most regions, assuming that all individuals are initially susceptible. The second factor is estimated from the inferred sigmoid curve for *R*_0_*_,t_*. The third factor is estimated assuming that case detection is followed by strict isolation, such that a detected case stops transmitting and the generation time is effectively truncated (Fig. 4A). This assumption is compatible with evidence that generation times are shortened by case isolation (17,18). With our set of parameters, the reduction owing to detection and isolation is approximately 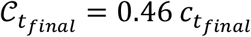. That is, on average detection and isolation only prevents 46% of transmission of a detected individual. The resulting reduction in transmission caused by detection and isolation is typically small (even under the conservative assumption that all detected individuals are perfectly isolated) because a small fraction of infected individuals is detected, and because individuals are detected a few days after symptoms when about half of the transmission already occurred (5,6).

To estimate the number of deaths averted by social distancing from the beginning of the epidemic to May 8^th^, we simulated the epidemic in the absence of social distancing measures, i.e. when transmissibility remains constant at its inferred initial value. The difference between the simulated number of deaths and the true reported number of deaths is the number of deaths averted. The reductions in transmissibility fom the beginning of the epidemic to May 8^th^ avoided in total across these regions 9.8 millions deaths, and of the order of ten thousands to one million deaths per country. Brazil, Mexico and large European countries (France, Germany, Italy, Spain, United Kingdom) avoided 5 to 8 × 10^5^ deaths. A previous study of 11 European countries reported figures similar to ours (5 to 7 × 10^5^ deaths deaths deaths avoided in the five aforementioneds countries (11)).

### Mobility as a correlate of transmission

the inferred time-varying transmissibility correlated with indicators of mobility. This has been evidenced in other studies in the USA (19,20). Precisely, we use the Google COVID-19 Community Mobility Reports which record the presence of individuals each day at six types of location: grocery & pharmacy, parks, transit stations, retail & recreation, residential, and workplaces, for most regions studied here (with the exception of Algeria, the Hubei province in China, Iran, Morocco, Russia, Ukraine). We used a multivariate linear mixed model to fit the reduction in transmissibility compared to baseline, as a function of the reduction in mobility indicator compared to baseline, in each country, at each day. The multivariate model corrects for the correlations between the different mobility indicators. The model includes mobility as a fixed effet, and region as a random effect affecting both the intercept and the slope of the relation. Interestingly, we found a correlation between transmissibility and all indicators, and particularly mobility in transit stations (public transport hubs such as subway, bus, and train stations). The model had a coefficient of determination of 93%. A reduction in the mobility in transit stations compared to baseline approximately translated in the same reduction in transmissibility (Table 1). This correlation was found in the whole dataset, in the subset of European countries, and in the subset of USA states.

**Table 1:** Regression coefficients fo the temporal relationship between transmissibility and each of the mobility indicators, in the full dataset, the USA states, and European countries. 95% confidence intervals indicated in parenthesis.Consistent and significant effects are highlighted.

| Dataset / model | Grocery / pharmacy | Parks | Transit stations | Retail & recreation | Residential | Workplaces |
| --- | --- | --- | --- | --- | --- | --- |
| full | -0.06 [-0.27 ; 0.15] | **-0.14** [-0.17 ; -0.1] | **1.02** [0.82 ; 1.21] | 0.13 [-0.08 ; 0.34] | -0.26 [-0.46 ; -0.06] | -0.02 [-0.15 ; 0.11] |
| USA states | -0.27 [-0.39 ; -0.15] | **-0.2** [-0.25 ; -0.15] | **1.31** [0.95 ; 1.64] | 0.17 [-0.12 ; 0.44] | 0.02 [-0.51 ; 0.53] | -0.26 [-0.52 ; 0.02] |
| Europe | 0.33 [-0.03 ; 0.63] | **-0.12** [-0.16 ; -0.09] | **1.02** [0.8 ; 1.24] | -0.14 [-0.43 ; 0.16] | -0.1 [-0.34 ; 0.15] | 0.36 [0.2 ; 0.53] |

**Table 2:** Summary of model parameters

| Parameter | Symbol | Value | Reference |
| --- | --- | --- | --- |
| Distribution of generation time | $w(\tau)$ | Log-normal(1.77, 0.888)<br>Mean 7 days<br>SD 4.5 days | (9,17,32) |
| Distribution of time from infection to symptom onset | - | Log-normal(1.518, 0.472) | (14) |
| Distribution of time from infection to death | $x(\tau)$ | Calculated by convoluting distribution from infection to onset and from onset to death. The latter is Gamma(5, 0.25) | (9) |
| Distribution of time from infection to detection | $y(\tau)$ | Calculated by convoluting distribution from infection to onset and from onset to detection | (14) |
| Infection fatality ratio | $d$ | Depends on the age structure of the country, around 1% on average | (8) |
| Probability of detection | $c_t$ | Inferred | - |
| Transmissibility | $R_{0,t}$ | Inferred | - |

### Relationship between probability of detection and intensity of testing

We last relate the time-varying probability of detection to the intensity of testing. First, we correlate the probability of detection (as of May 8^th^) with the number of tests performed by inhabitants *across regions*. We did so for 62 regions where test data were available. There was a strong correlation between probability of detection and number of tests per inhabitants (regression coefficient β = 161 per daily test per inhabitant, *p* = 4.0 × 10^-5^) (Fig. 5B).

**Figure 5:**
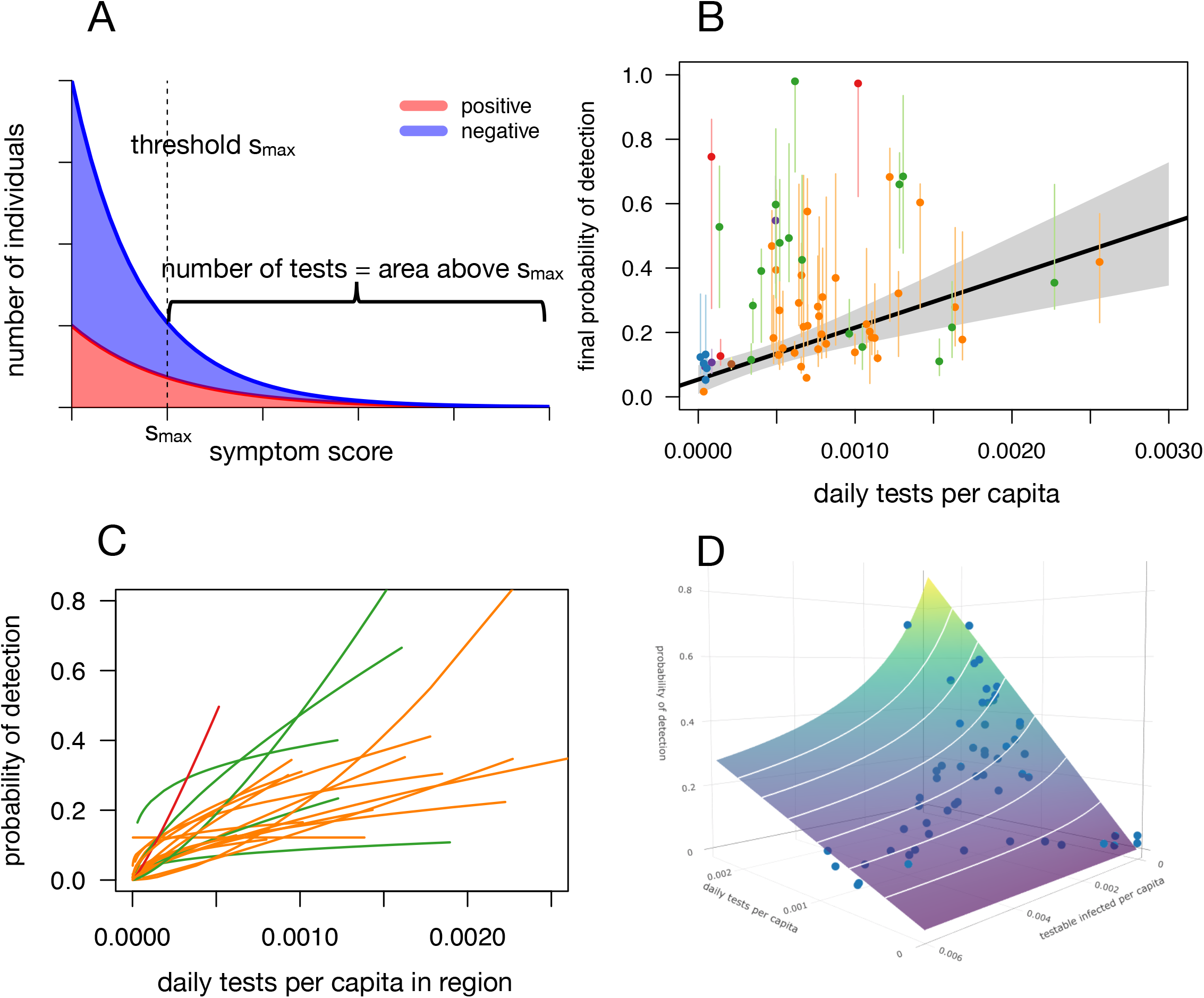
Relationship between probability of detection and number of tests. **Panel A** shows stacked distributions of the disease score for positive (red) and negative (blue) individuals. The fraction of positive individuals increases with the score. The number of tests performed is the area to the right of the threshold (vertical line). **Panel B** represents the final probability of detection as a function of number of daily tests per capita (over the 7 days preceding 8th May) for the 62 regions with available test data. **Panel** C shows the predicted root-function relationship between proportion of detected and daily tests for the 33 regions with sufficient available test data. **Panel D** shows the proportion of detected a function of daily tests and the number of testable infected presenting for a test, for the New York state (one of the high-prevalence states where the proportion detected declines with the number of infected as predicted at high prevalence).

Second, to examine further how the changing number of tests affects the probability of detection *within a region and across time*, we formulated a simple model of testing. The goal of this model is to relate within a region the number of tests conducted on a given day (called *T_t_*) with the inferred probability of detection on that day (*c_t_*). We assume that in the period when the incidence of infections is much higher than the number of tests, the decision to test individuals for SARS-CoV-2 is made on the basis of a set of symptoms and risk exposure defining a score. SARS-CoV-2 infected and uninfected individuals present two distinct distributions of this score, such that the probability that the individual is truly infected with SARS-CoV-2 increases with this score. Tests are prioritised on individuals with the highest score. This model thus reproduces the fact that the fraction of positive tests increases when tests are more limited compared to the number of infected individuals. For simplicity, we additionally assume that the score in infected and uninfected individuals follows exponential distributions with two distinct rates. Under this model, the probability of detection is given by the solution *c_t_* of:

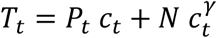

(Material and Methods). In this equation, the variable *T_t_* is the total number of tests conducted at day *t. P_t_* and *N* are the number of SARS-CoV-2 infected and non-infected individuals seeking care at day *t*, and who could potentially be tested if the number of tests available allows. *P_t_* is the time-delayed number of infected individuals given by 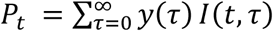 where *y*(*τ*) is the probability that an individual is detected *τ* days after infection (when it is detected), while *N* is assumed to be constant over the considered period. The parameter *γ > 1* describes the distribution of the symptom score in infected individuals relative to that in uninfected individuals. There is no closed form solution for the general solution *c_t_*, but when the distribution of the score is dominated by negative individuals, the probability of detection is approximately a root function of the number of tests:

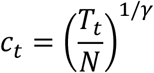

The probability of detection should thus generally increase sublinearly with the number of tests since *γ* > 1, and at best, should be proportional to the number of tests (when *γ =* 1). This is because tests are prioritised on individuals that are more likely to be infected; as the number of tests increases, the probability of positivity decreases. We also predict that in general, when the number of infected is large, the probability of detection decreases with the number of infected individuals (Material and Methods).

Both predictions were verified in data (Fig. 5). We inferred for each region the best-fitting pair of parameters (*N, γ*) to relate the inferred probability of detection *c_t_* to the number of tests *T_t_*, using both the approximated and the general model. We found that *γ* > 1 for most regions, implying a sublinear relationship as predicted (Fig. 5C). The general model where the probability of detection decreased with the number of testable infected individuals was a better fit only when the attack rate was high, for example in New York state (Fig. 5D, Supplementary Fig. 8).

## Discussion

We developed a discrete time renewal equation model to describe the dynamics of SARS-CoV-2 infections. We fitted this model to the daily cases and deaths in a large number of countries and states (together representing 4.2 billions individuals), with the following results:

i. Transmissibility declined in all 79 regions examined. The best-fit decline in transmissibility was often smooth, with the decline in transmissibility predating the date of the lockdown. This could be due to non-pharmaceutical interventions implemented before the full lockdown or other behavioural changes. However, the decline in transmissibility as of May 8^th^ was not enough to contain the epidemic in a number of regions.
ii. The probability of case detection increased, was on average 29% across regions as of May 8^th^, and very rarely above 50%.
iii. Epidemic control was achieved mainly through reductions in transmissibility brought about by social distancing. Case detection and isolation had a limited impact (Fig. 4B), even under the conservative assumption that case detection is followed by perfect isolation. Only a small proportion of cases are detected and about half of the transmission happens before symptom onset. We emphasise that in this period most testing was based on symptoms and not on past contacts with infected individuals. The build-up of immunity in infected individuals also had a very limited impact because the fraction of individuals infected remains small in all regions. Social distancing in the regions considered avoided almost 10 millions deaths from the beginning of the epidemic to May 8^th^.
iv. Transmissibility correlated with mobility indicators, and most notably with the presence of individuals in transit stations, both in Europe and in the USA.
v. The inferred probability of detection correlated with the number of tests per capita across regions. However, increasing the number of tests does not proportionally increase the probability of detection. This is explained by the fact that tests are prioritised on individuals most likely to be infected.

Our model and inference rely on several assumptions. First of all, we describe transmission dynamics within a simplified model that does not take into account age structure or household structure. These forms of structure may be weak enough that they can be neglected when describing the overall epidemic trajectory (21). Second, to infer jointly the time-varying transmissibility and probability of detection within a dynamical model, we assumed the temporal change took specific sigmoid functional forms. This differs from other approaches which estimate daily transmissibility as the incidence at a given day divided by past incidence weighted by the distribution of the generation time (22,23). These alternative approaches are more flexible in that they can infer any pattern of time-varying transmissibility. However, they cannot account exactly for the delay in case reporting, and can be very sensitive to noise in the data (22). Fitting a dynamical model with imposed functional forms for transmissibility and probability of detection reduces the sensitivity of inference to noise in the data. Third, and most importantly, inference relies on daily deaths and cases. Deaths are assumed to be perfectly reported. Cases are assumed to be partially reported with a time-varying detection probability. The inferred absolute value of the probability of detection of course strongly relies on the assumed IFR at around 1% on average (and tuned to the specific age structure of each region considered). The approach was validated in a number of regions where systematic test or seroprevalence surveys were conducted (Fig. 1). It is possible that in some of the other regions examined the number of deaths was greatly under-reported, in which case the true number of infected would be much higher than predicted, and the probability of case detection much smaller. However this should not affect the temporal trends in transmissibility or probability of detection, provided that under-reporting is constant in time. Other emerging seroprevalence surveys will give more information on the IFR (or death under-reporting) across regions, but it is notable that the early estimate of IFR in mainland China (8) already allow good predictions (Fig. 1). Lastly, our framework does not take into account the possibility that the IFR changes in time. Such temporal variation in IFR could be caused by overwhelmed health systems (increasing IFR) of better social distancing in at-risk groups (decreasing IFR).

Our method has several advantages. The discrete-time renewal equation makes the minimal assumptions that the transmissibility of an infected individual depends on the age of infection. It allows arbitrary distributions of the generation time, and arbitrary delays between infection and case detection, and infection and death. The distributions of these delays determines the dynamics of the changes in number of cases and deaths following a change in transmissibility. Parameters can be inferred using multiple time series, improving the precision of inference.

The daily cases, although dependent on the number of tests available, give an earlier signal of changes in transmissibility than the daily deaths, and suffer less from stochastic effects. The method allows different transmissibility for detected cases (here assumed to be zero, i.e. perfect isolation after detection). This is particularly relevant for accurate inference of transmissibility, as non-pharmaceutical interventions shorten the serial interval (fig. 4A) (18). Lastly, the framework quantifies the immunity acquired by infected individuals.

The probability of detection as a function of time in different countries was computed by different means in another study (24). Their statistical approach was based on estimating the case fatality ratio (CFR) adjusted for the delay between infection and deaths, and comparing with the baseline infection fatality ratio estimated in other studies that account for under-reporting (assumed to be 1.4% in their case). Their statistical method allows inferring arbitrary temporal variations in the probability of detection. However, it does not explicitly model the dynamics of transmission. It is unclear how the changing age-of-infection structure of the population upon reductions in transmission will affect the relationship between daily number of deaths and past number of cases, hence the inferred probability of detection, in their approach.

We found that tests detected only a small proportion of cases. Furthermore, increasing the number of tests does not proportionally increase the proportion of detected individuals. As a consequence of the typically small probability of detection, together with the fact that a large part of transmission already occurred at test result, tests followed by isolation of positive cases had very little impact on transmission, and were not sufficient by themselves to control an epidemic with a basic reproductive number of 3 or more. We assumed that individuals isolate only at the date of the test result (a few days after symptom onset). Assuming that symptomatic individuals isolate at symptom onset would not change much our quantitative results. Our model assumes an exponential distribution of the score underpinning the decision to refer an individual to a SARS-CoV-2 test or not. This distribution could be linked to more precise data if individuals tested in priority are those most likely to be infected, and if the decision to test is based on a defined set of variables describing symptoms, or risk exposure. For example, a logistic regression of infection status vs. symptoms (as in (25)) would define a score for each individual based on a linear combination of these symptoms. The probability of infection would increase with this score, and the right tail of the distribution of this score (including the individuals most likely to be infected) could resemble an exponential distribution.

Our model is primarily concerned with the first few months of the epidemic where in most regions contact tracing could no longer be practically implemented. More widespread contact tracing could improve the relationship between probability of detection and number of tests, as contacts of positive cases may have a 5-10% chance of being positive, up to 10-15% for household contacts (17,26,27). Furthermore, contacts could self-isolate before the onset of symptoms (5,28). For these two reasons, contact-tracing and testing is a more efficient way to control the epidemic than symptom-based testing. Thus, if the capacity to trace contacts is limited, the epidemic may be out control as soon as the daily incidence is too large to trace a good fraction of contacts. This pleads for the use of digitical contact tracing apps and/or rapid implementation of additionnal social distancing measures when incidence increases (5).

Lastly, the inferred time-varying transmissibility correlated with mobility indicators (20,29). More precisely, within a multivariate framework we found that the mobility in transit stations was the most highly correlated with transmissibility, a pattern consistent in European countries and the USA (Table 1), and with a regression coefficient close to 1 (a given reduction in mobility corresponds to an equivalent reduction in transmission). The mobility in transit stations could be a general indicator of economic / social activity resulting in more transmission. Public transports could also be a common context of transmission. In support of our finding, individual use of public transport in Maryland was strongly associated with SARS-CoV-2 positivity (30).

In conclusion, we developed a framework to estimate time-varying transmissibility and probability of detection from daily cases and deaths in a large number of countries and regions. In the first few months of 2020, control of the epidemic was achieved mostly by reductions in transmissibility, which avoided 10 millions deaths in these 79 regions (representing more than half of the world’s population), while case detection and isolation comparatively had a much smaller effect.

## Methods

### Deterministic transmission dynamics

To model transmission dynamics, we use a discretised version of the renewal equation (e.g. (11)). We follow the dynamics of the number of individuals infected at day *t* who were infected *τ* days ago, and have not yet been detected and isolated, called *I*(*t*, *τ*). The transmission dynamics are given by the system of recurrence equations:

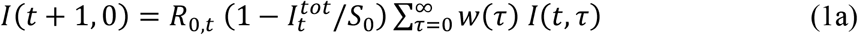

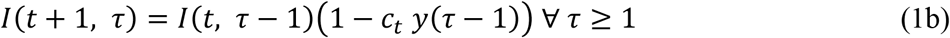

The first equation represents transmission to new susceptible individuals giving rise to infected individuals with age of infection 0. The parameter *R*_0_,*_t_* reflects transmissibility, and is the basic reproduction number (in the absence of interventions, and when the population is fully suscpetibe, i.e. 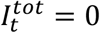). The factor *w*(*τ*) is the fraction of transmission that occurs at age of infection *τ*, where 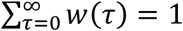. Thus *w*(*τ*) represents the distribution of the generation time of the virus. The infectiousness profile of the virus is linked with the generation time distribution through *β*(*τ*) = *R*_0,_*_t_ w*(*τ*). Transmission is reduced by a factor 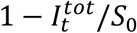 by population immunity, where *S*_0_ is the initial number of susceptible individuals in the region, assumed to be the total population size. The variable 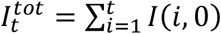 is the total number of individuals already infected and assumed to be fully immune at time *t*. The instantaenous reproduction number that accounts for population immunity but not for case isolation is = 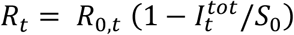.

The second equation (1b) represents the dynamics of individuals infected in the past. Individuals infected *τ* − 1 days ago are now of age of infection *τ*, provided they were not detected and isolated. An infected individual is detected with time-varying probability *c_t_*, and the probability that an individual is detected at age *τ* (when it is detected) is given by *y*(*τ*), with 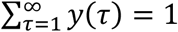. An individual who is detected is removed from the pool of individuals that contribute to further transmission of the disease. The total number of cases detected at day *t* is thus:

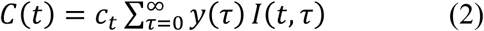

And the number of detected individuals who were infected *τ* days ago changes as:

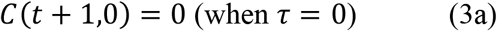

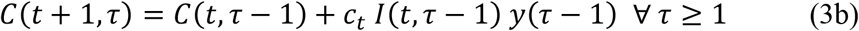

The total number of infected individuals, be they undetected or detected, that we may call *A*(*t, τ*) *= I*(*t, τ*) *+ C*(*t, τ*), follows the equations:

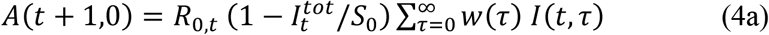

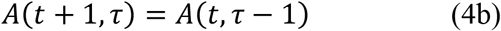

The fact that incidence (in the first equation) only depends on undetected cases *I*(*t, τ*) emerges from the assumption that detected individuals *C*(*t, τ*) do not transmit.

While in the absence of testing and isolation, the infectiousness profile is given by *β*(*τ*) *= R_0_,_t_ w*(*τ*) (with 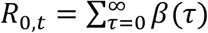, detection and isolation truncates the infectiousness profile at the time of detection *t_d_* (Fig. 4A) with probability *c_t_ y*(*t_d_*) where *t_d_* is the time of detection. In other words, the effective infectiousness profile is the mixture distribution:

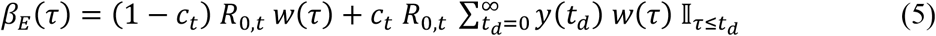

where 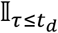 is an indicator variable equal to 1 when *τ ≤ t_d_*, and 0 otherwise.

### Probability of dying and time to death given infection

The probability that an infected individual dies is the infection fatality ratio (IFR) denoted *d*, assumed to be constant over time. The probability of dying *τ* days after infection, given that one dies, is given by *x*(*τ*). The mean number of deceased individuals at day *t* is then given by:

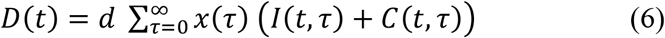

As death typically occurs at a time when the infected individual does not transmit any longer, and the probability of dying is small (of the order of 1%), we neglect the impact of death on transmission.

### Effects of detection and isolation, change in transmissibility and immunity on transmission

We call “effective reproduction number” 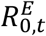 the instantaneous reproduction number taking into account immunity and case isolation is given by (see also (31)):

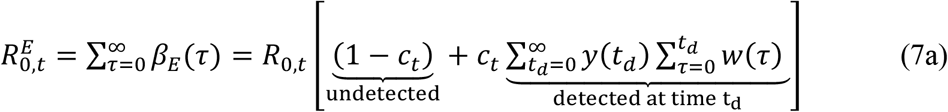

For example, an individual detected at day 0 only infects *R_0,t_ w*(0) individuals on average. Equation (6a) can be rewritten as:

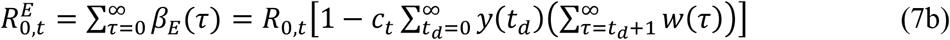

Thus, the effective reproduction number 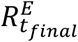 on the 8^th^ of May (*t_final_*), including the impacts of detection and isolation and immunity may be written as the product of the initial basic reproduction number, times three factors that all reduce transmission:

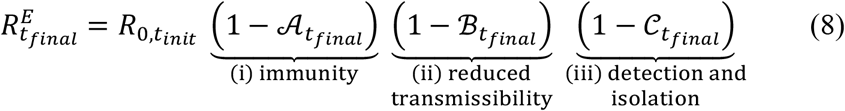

With 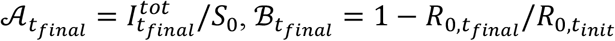, and 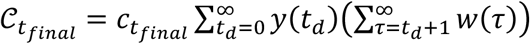.

### Parameter estimates

We fix the distributions of the generation time *w*(*τ*), the distribution of time from infection to death *x*(*τ*), the distribution from infection to detection *y*(*τ*), and the infection fatality ratio *d* to values estimated previously.

#### Generation time

We assume the generation time is lognormally distributed with mean 7 days and standard deviation 4.5 days (9) (Supplementary Fig. 4). This is the generation time when the infected individual is not tested. A positive test is assumed to be followed by perfect isolation of the infected individual and interruption of transmission. This effectively truncates the distribution of generation time (Fig. 4A). Two factors make estimation of this generation time difficult: first, the generation time, the time from an infection to another infection, is often approximated by the serial interval, the time between symptom onset in an infector and symptom onset in the infectee. These two quantities have the same mean, but the variance of the generation time should in general be smaller than that of the serial interval (33). Second, measuring the serial interval requires to identify infectees and their infectors. The fact that the infector needs to be identified could bias the serial interval towards lower values. For example, in a large study in the Shenzhen province in China, the serial interval had mean 6.3 days overall and 8.1 days if the infector was isolated more than two days after infection (17). Thus, in settings where most infections are undocumented, the typical serial interval (and generation time) may be longer than that estimated in other work (e.g. mean 5 days in (5)), motivating the mean of 7 days chosen here.

Note that the chosen serial interval distribution affects the absolute value of the basic reproduction number, but does not affect either the inferred temporal trend in basic reproduction number or the absolute value of the probability of detection.

#### Time from infection to detection

The time from symptom onset to case detection was inferred from published data on 150 cases from various countries (14). We used the time between the midpoint date of symptom onset and the midpoint date of case detection. We excluded 31 cases for which the date of case detection was not available or there was very large uncertainty on the date of symptom onset. We inferred that the time from symptom onset to detection was gamma-distributed with mean 2.2 days [95% CI 1.6-3.2] and SD 2.7 days [2.0-3.8] (shape 0.69 [0.55-0.82] and rate 3.2 [2.5, 4.5]). The fit of a Weibull distribution was comparable to that of the gamma (Supplementary Fig. 5).

The distribution of time from infection to detection was computed from the convolution of the distribution of time from infection to symptom onset (14), and our inferred distribution of time from symptom onset to case detection, assuming independence of the two times. The distribution of time from infection to symptom onset has mean 5 to 6 days (Supplementary Fig. 6).

#### Time from infection to death

The distribution of the time from infection to death was estimated using data from 41 patients in Wuhan analysed elsewhere (12). The time from symptom onset to death was gamma-distributed with a mean of 20 days and a standard deviation of 10 days. This estimate is close to that of other studies (24 deceased cases from mainland China, mean and SD of time from onset to deaths 18.8 / 8.5 days (8); 34 deceased cases from mainland China, mean and SD 20.2 / 11.6 days (34).

#### Infection fatality ratio

For each region studied, we computed an overall infection fatality ratio that takes into account the age pyramid of the country. To this end, we used the infection fatality ratio (IFR) estimated in nine age classes, [0, 9], [10, 19], etc., [80+] in mainland China (8). Other estimates similarly stratified by age, for mainland China and for France, are very similar (Supplementary Fig. 1). The IFR climbs from close to 0% in 0 to 39 years old, up to 5 to 10% in individuals aged 80 years old or more.

### Likelihood method

To fit the model and infer transmission and case detection parameters, we use data on the ^1^ number of confirmed cases over time and the number of deaths over time in 79 states and countries from different public sources detailed below. We include all states and countries that had a daily incidence of 10 deaths or more at least once as of 23th April. As we want to estimate the impact of sudden social distancing measures in an essentially uncontrolled epidemic, we exclude South Korea and Japan from the analysis. In these two countries, SARS-CoV-2 was introduced earlier and strong control measures including social distancing and contact tracing were immediately in place.

The likelihood of the model is defined as in other epidemiological studies (7,11). Simulating the deterministic model gives the expected number of detected cases *C*(*t*) and deaths *D*(*t*) at time *t* as a function of model parameters. We assume that the probability to observe a certain number of cases (resp. deaths) in the data at day *t* is the density of a negative binomial distribution with mean given by the theoretical predictions for cases (resp. deaths), and dispersion parameters that we infer. The overall likelihood is the product of these probabilities over all days. For the number of deaths, we include the period from the first day to the last day when at least 1 death and 5 cases were recorded. For the number of cases, we include the period from the first day to the last day when at least 5 cases were recorded. We offset the simulation time such that the date when 5 deaths are reached in the simulations becomes the date when 5 deaths are reached in the data. At this date, the number of infected is large enough that the underlying dynamics should largely be deterministic.

We mainly estimate the time-changing transmissibility *R*_0_,*_t_* and the time-changing probability of detection *c_t_*.

For the time-changing transmissibility, we fit two functional forms. First we assume that *R*_0_,*_t_* is a step function with a sharp transition from a high pre-control value to a low post-control value, at a fixed date *t_control_* corresponding to the date of implementation of the control measure:

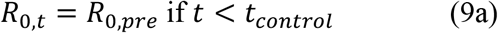

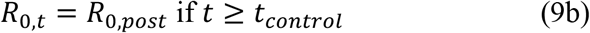

For the sharp change in transmissibility, we infer the two values *R*_0_,*_pre_* and *R*_0_,*_post_*. Furthermore, to investigate the possibility that transmissibility changed in a more gradual way, we assume *R*_0_,*_t_* is a smooth declining sigmoid function:

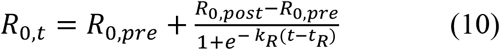

Where *R*_0_,*_pre_* is the basic reproductive number before social distancing measures, *R*_0_,_post_ is the basic reproductive number after social distancing, *k_R_* is the steepness of the logistic curve, and *t_R_* is the time when the basic reproductive number is intermediate between *R*_0_,*_pre_* and *R*_0_,*_post_*. The step function is a special case of the logistic when *k* is large and *t_R_* = *t_lockdown_*. For the smooth change in transmissibility, we infer the two values *R*_0_,*_pre_* and *R*_0_,*_post_*, the steepness and the time *t_R_*.

For the time-changing detection probability, we assume an increasing logistic function:

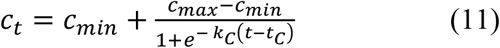

We infer the four parameters *c_min_*, *c_max_*, and *t_c_*. Note that we constrain the parameter *c_min_*, the initial probability of detection, to be small, in [0.0001, 0.001]. We fit three models: (i) a model based on death data only with the step function of transmissibility, (ii) a model based on death and case data with the step transmissibility function; and (iii) a model based on death and case data with the smooth transmissibility function. These three models are fitted by maximum likelihood. We first find an optimal likelihood value by 50 iterations of the Nelder-Mead algorithm starting from different initial parameters. We then run a Markov chain Monte Carlo (MCMC) sampling of the likelihood function with bounded parameters (equivalent to uniform priors for all parameters in a Bayesian framework). We let the chain run for *10^6^* steps and record the parameter values from 2 × 10^5^ to *10^6^* steps. This sample is used both for maximum likelihood parameters (if a better parameter set is found than with the Nelder-Mead algorithm) and for confidence intervals.

### Symptom-based test model

#### The model

We relate the fraction of infected individuals detected to the number of daily RT-PCR tests performed and the incidence of infection. Each day, the testable individuals seeking care are composed of two populations:

- SARS-CoV-2 infected individuals. The number of such individuals is time-varying and is denoted by 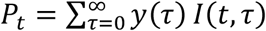 where *y*(*τ*) is the probability that an individual is detected at age of infection *τ* (given that it is detected) (Fig. 5A).

- Non-SARS-CoV-2 infected individuals. The number of such individuals is assumed to be constant and is denoted by *N*. We acknowledge that a more complete model would allow for this number to vary in time, for example to account for seasonal infections by respiratory diseases like influenza or seasonal coronavirus that may contribute to the pool of testable individuals.

We assume that contexts in which we apply our model are characterized by a number of tests smaller than the number of testable individuals, *T_t_ < P_t_ + N* where *T_t_* is the number of tests available at time t. Thus the *T_t_* tests are prioritised on the subset of individuals most likely to be infected by SARS-CoV-2. Individuals seeking care are characterised by a score such that the probability of SARS-COV-2 infection increases with the score. Given the limited number of tests available each day, a threshold score is defined and tests are performed only for patients above this score. Formally, denoting by 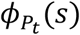 and *ϕ_N_*(*s*) the distribution of the score *s* in infected and uninfected individuals, the (time-varying) threshold score *s_max_* is the solution of:

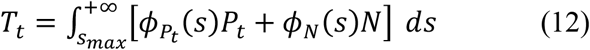

In the absence of detailed information on the choice of individuals to test in different regions at different stages of the pandemic, we further assume for simplicity that the scores are distributed exponentially. We set the rate of the exponential distribution 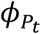 to 1 without loss of generality, and we denote *γ* > 1 the rate of *ϕ_N_*:

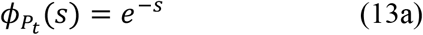

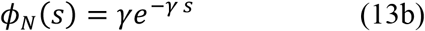

The fact that *γ* > 1 guarantees that the probability that an individual is positive increases with the score. Plugging the distributions (13) in the implicit formula to define the threshold score *s_max_* (12) yields:

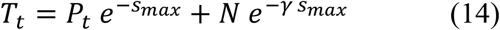

The probability of detection *c_t_*, defined as the ratio between positive tests results and the number of testable infected individuals *P_t_*, is the area of the *ϕP_t_* distribution above the threshold

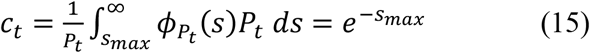

Replacing with equation (15) in equation (14), we find that *c_t_* is the solution of:

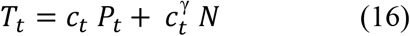

This generally defines an implicit function *c_t_*(*r_t_*, *P_t_*) of the number of testable infected at day *t*, *P_t_* and the number of available tests *T_t_*. We can simplify this general solution in two ways. First, in the limit when the number of infected *P_t_* is much smaller than the number of uninfected *N*, and γ is not too large, the probability of detection is:

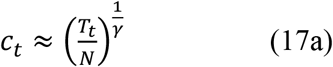

That is, the probability of detection increases as a root function of the normalised number of tests. In general, when the number of testable infected individuals *P_t_* is not negligible, the solution *c_t_* of equation (16) decreases with *P_t_*. When the number of testable infected individuals *P_t_* is small, the solution (17a) can be better approximated by:

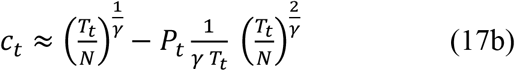

In this approximation the probability of detection decreases linearly with the number of infected *P_t_*.

#### Parameter inference

We verify the model predictions using the inferred probability of detection *c_t_* together with data on the daily number of tests *T_t_*, and the number of testable infected individuals *P_t_* inferred from the dynamical model in different regions. We used the *nls* method from the *stats* package in the software R (35).

First, we use the general solution of equation (16). This solution is a non-linear function *c_t_*(*T_t_, P_t_*) with parameters *N* and *γ*. We infer the parameters *N* and *γ* by minimizing the mean square error between the inferred *c_t_* and the prediction. In most cases (except, notably New York and New Jersey states) the coefficient of determination was as good with the simplification of the model where *c_t_* is approximated as a root function of *T_t_* only (equation(17a)) (Supplementary Fig. 8). The general solution improved the fit all the more than the the attack rate was larger, as predicted by the model (Supplementary Fig. 8).

## Data Availability

Code and data is available on https://github.com/FrancoisBlanquart/covid_model

## Data sources

### Epidemiological data

For France, we used data from OpenCOVID19 available at https://github.com/opencovid19-fr/data. This website curates data from *Agence nationale de santé publique*, the French governmental public health agency.

For Italy, we used data from the Civil Protection Department (Dipartimento della Protezione Civile), available at https://github.com/pcm-dpc/COVID-19. This data includes daily cases and deaths, and daily number of tests.

For other European countries, we used data from the European Center for Disease Control. (ECDC) available at https://opendata.ecdc.europa.eu/covid19/casedistribution/

For American states, we used data from the COVID Tracking Project that compiles data from American official sources, available at https://covidtracking.com/api/v1/states/daily.csv. This data includes daily cases and deaths, and daily number of tests.

For other countries, we used data from the Center for Systems Science and Engineering (CSSE) at Johns Hopkins University (JHU), available at https://github.com/CSSEGISandData/COVID-19.

Daily number of tests data for regions other than Italy and American state were compiled from Our World in Data at https://covid.ourworldindata.org/data/owid-covid-data.csv

We considered test data only for regions for which the number of tests was strictly superior to the number of cases recorded for at least 80% of the days. The reported number of tests are sometimes exactly equal to the number of cases that day, indicating that negative tests are not reported. Since we ignore whether negative tests are not reported or reported at a later date (as sometimes suggeted by a peak in the number of reported tests a few days after), we exclude these datapoints and exclude regions where this artefact is often observed.

### Age structure data

We collected data on the number of individuals in age categories 0-9, 10-19,…, 80+, for different states and countries, from the following sources:

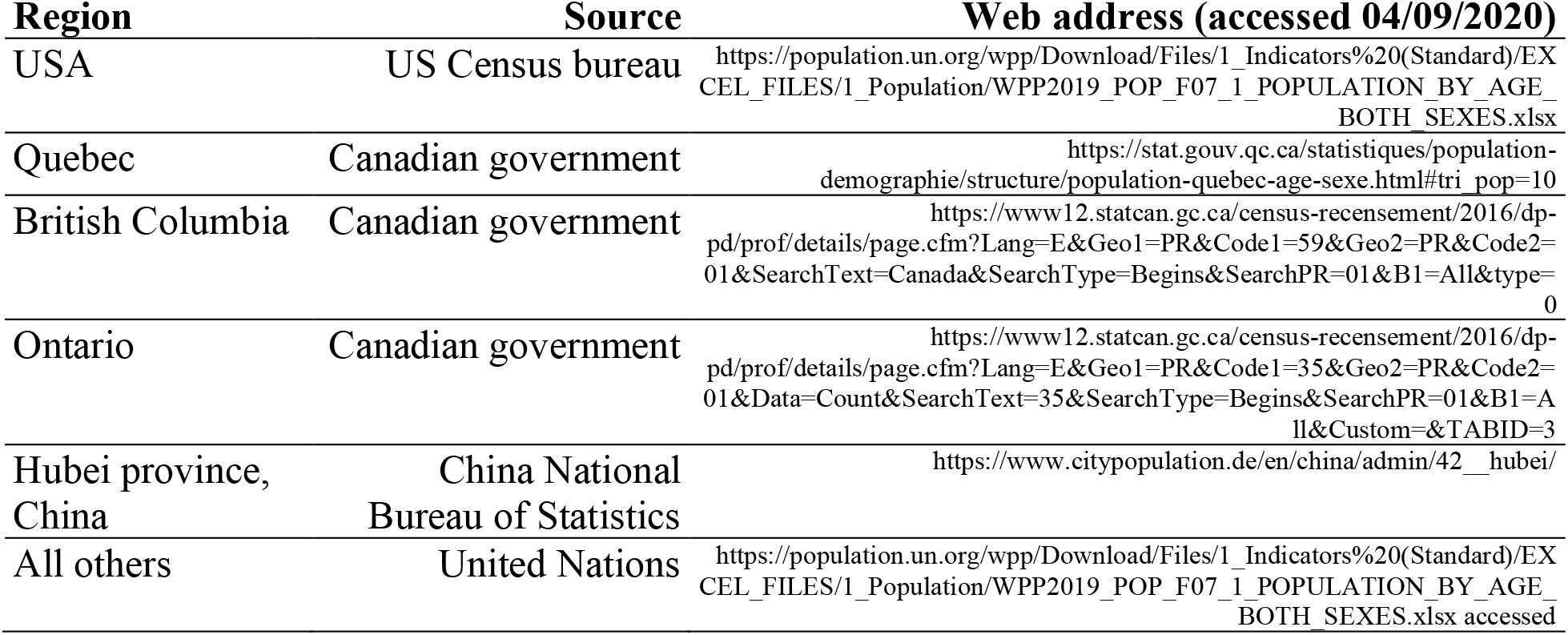

### Mobility data

We used Google mobility data available at https://www.google.com/covid19/mobility/

## Data availability

Code and data is available on https://github.com/FrancoisBlanquart/covid_model

## Acknowledgements

We thank Florence Débarre and Chris Wymant for helpful comments. We thank the many people involved in the collection and curation of the epidemiological data that we use. F.B. was supported by a Momentum grant from the CNRS. A.B was supported by a scholarship from Ecole Polytechnique.

## Supplementary Information for

**Supplementary Table 1:** table of systematic test or seroprevalence studies used for figure 1

| Location | PCR / sero test | Time interval | Prevalence (N) | Reference |
| --- | --- | --- | --- | --- |
| Austria | PCR | 4-5 April | 0.33% (1544) | https://unctad.org/en/PublicationsLibrary/CS_TD_COVID19_c06_Austria_en.pdf |
| Brazil | serological | 14-21 May | 1.4% (25,025) | (1) |
| New York state | serological | 20 April - 1 May | 12.3% (15,103) | https://www.governor.ny.gov/news/amid-ongoing-covid-19-pandemic-governor-cuomo-announces-results-completed-antibody-testing |
| Netherlands | serological | 1-15 April | 2.7% (7,361) | (2) |
| Denmark | serological | 6-17 April | 1.7% (9,496) | (3) |
| Spain | serological | 27 April - early May | 5% (60,983) | (4) |
| Belgium (1) | serological | 30 March | 2.1% (1,300) | https://www.sciensano.be/fr/coin-presse/43-de-la-population-belge-a-developpe-des-anticorps-contre-le-coronavirus |
| Belgium (2) | serological | 14 April | 4.1% (3,000) |  |
| Czechia | serological | 23 April – 1 May | 0.4% (26,549) | https://koronavirus.mzcr.cz/infekce-covid-19-prosla-ceskou-populaci-velmi-mirne-podobne-jako-v-okolnich-zemich/ |

**Supplementary Figure 1:**
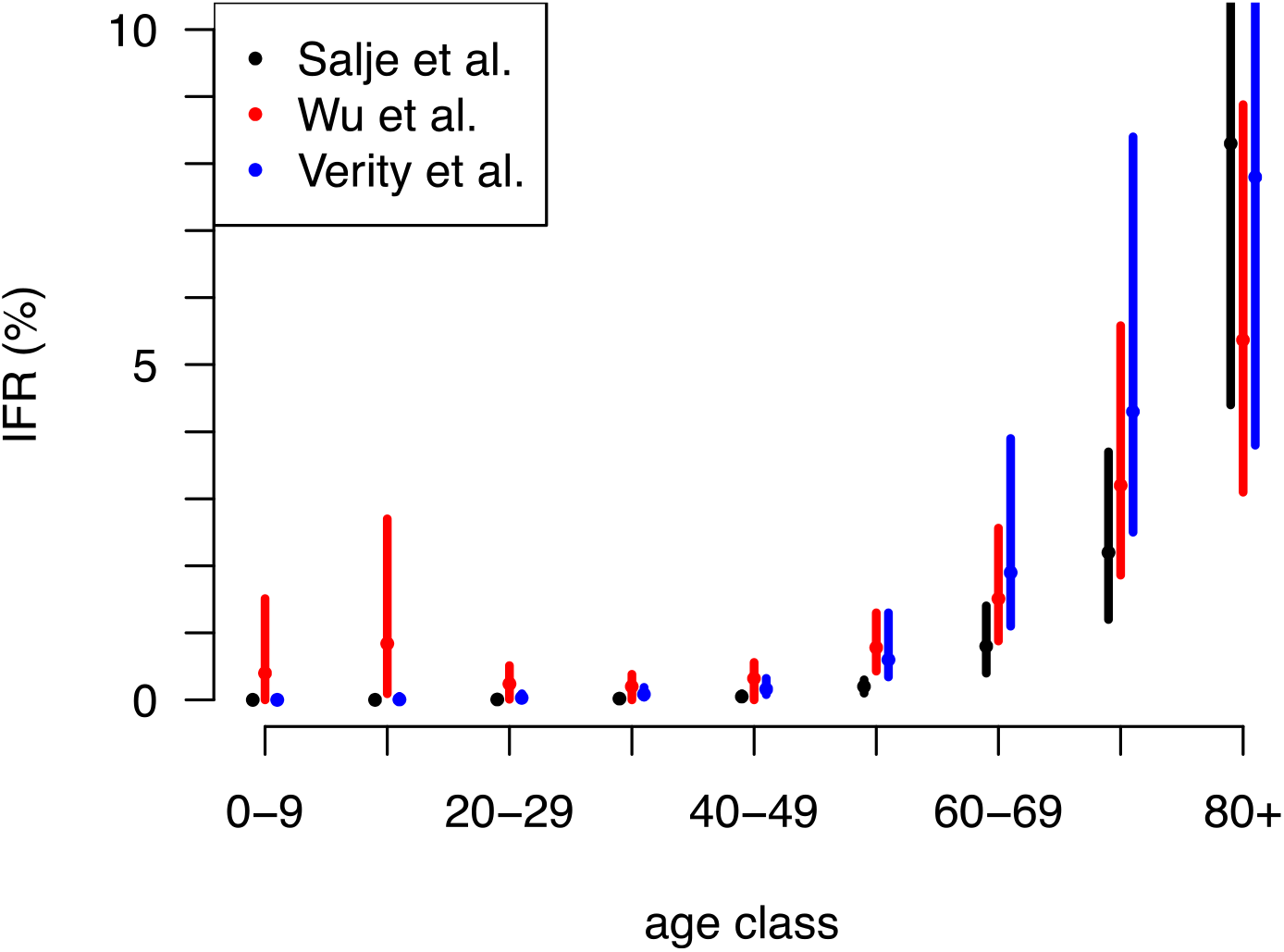
infection fatality ratio in %, with 95% confidence intervals, as a function of the age class, as estimated in three studies (5-7).

**Supplementary Figure 2:**
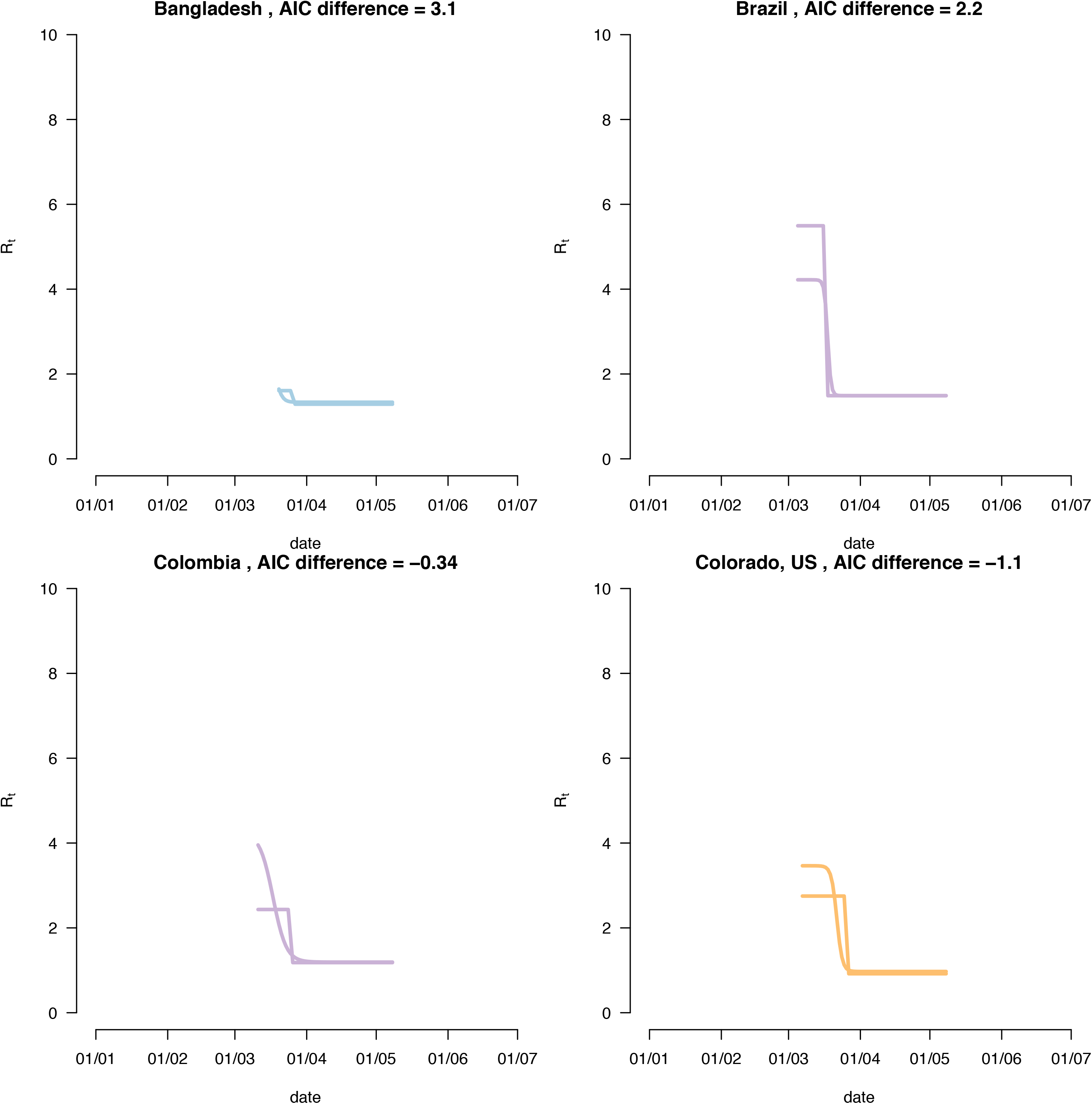
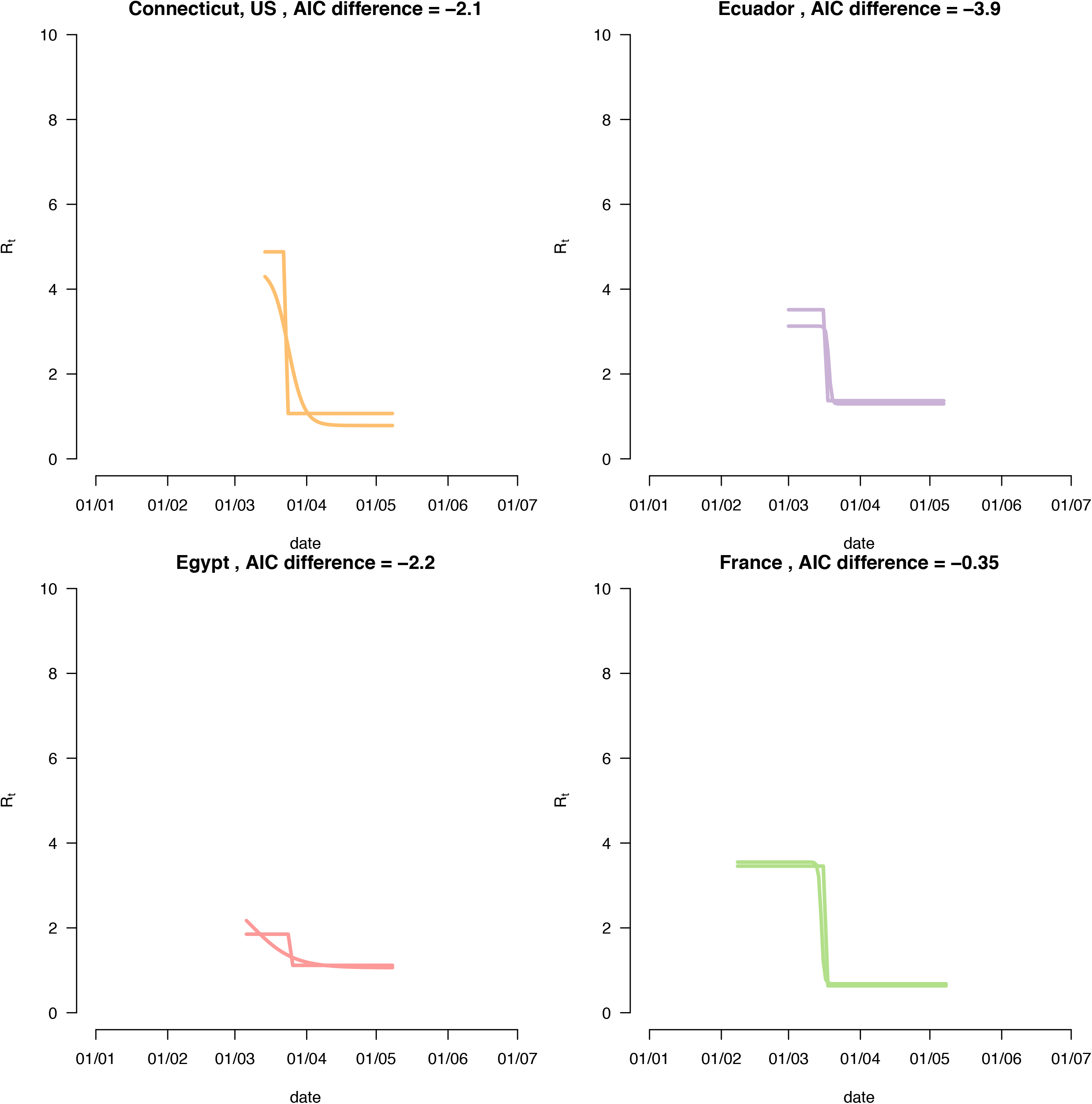
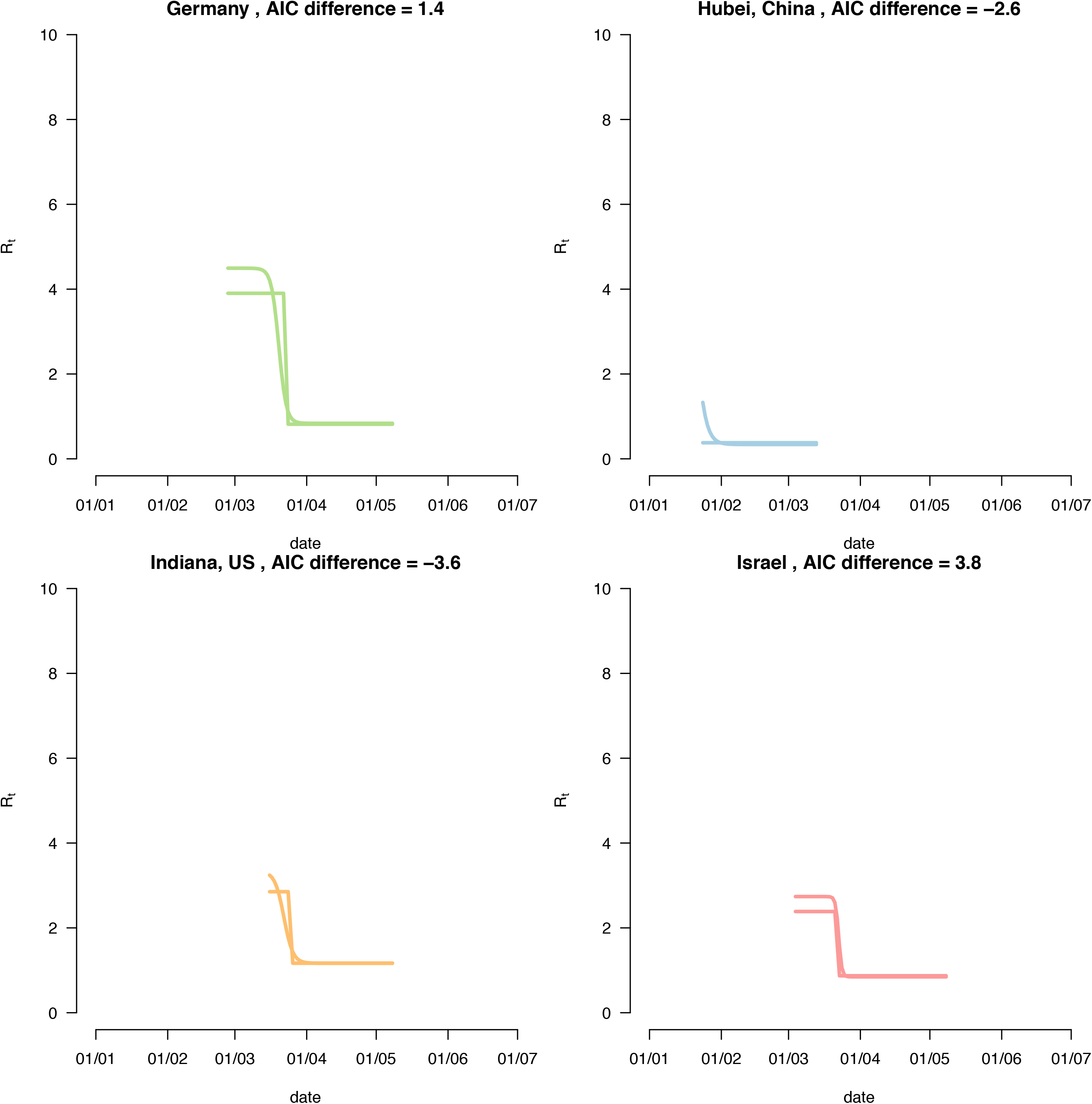
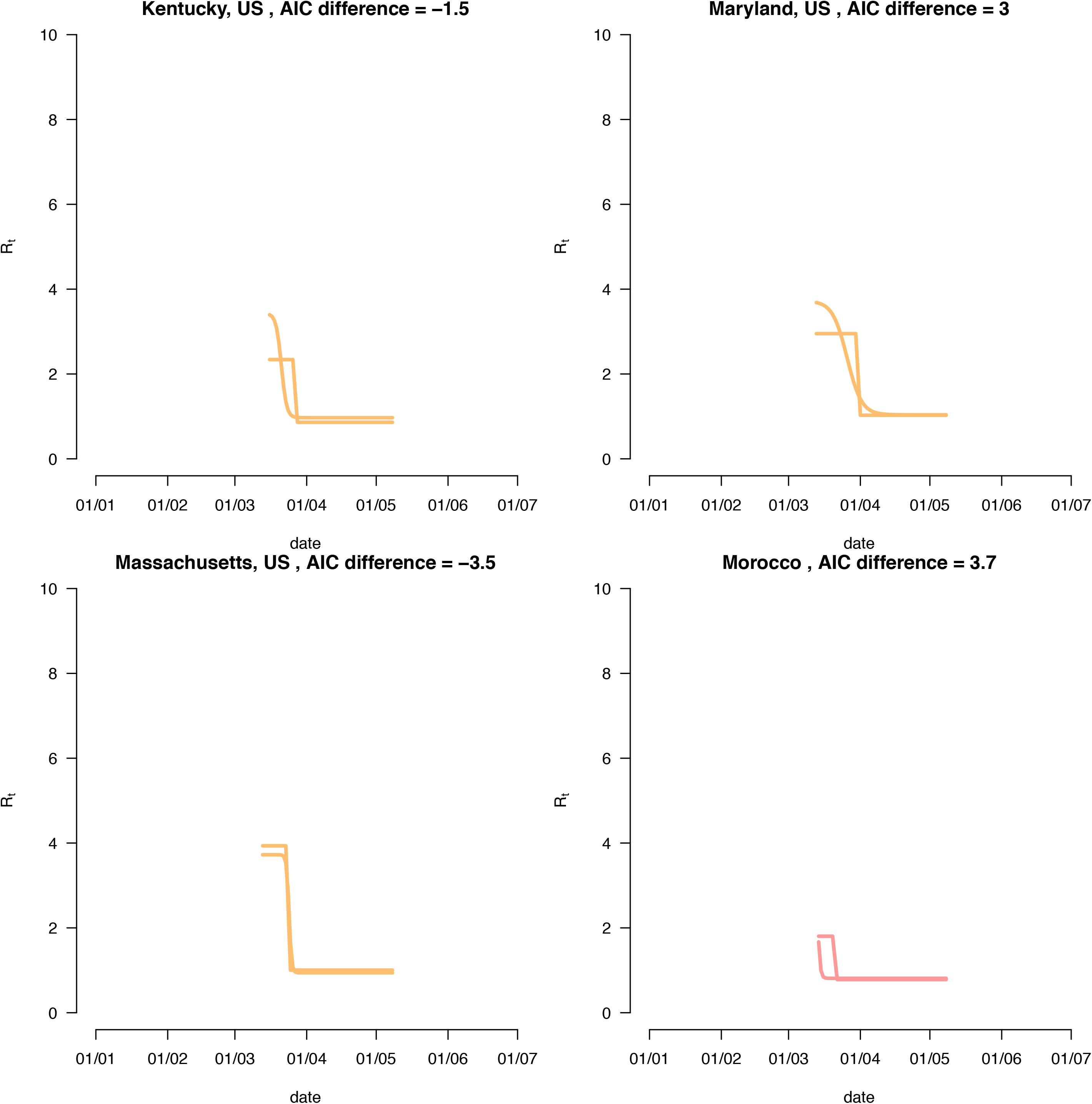
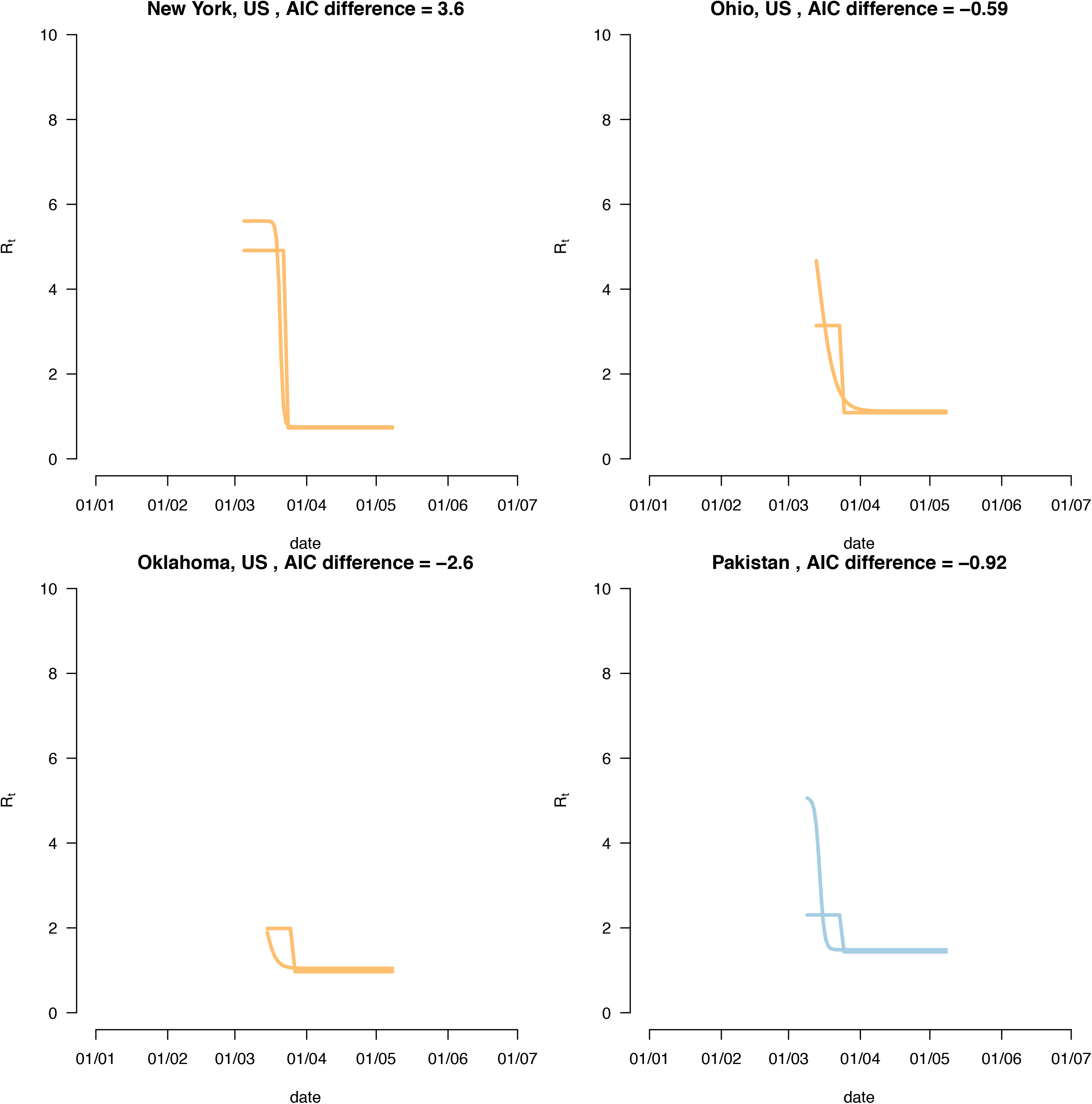
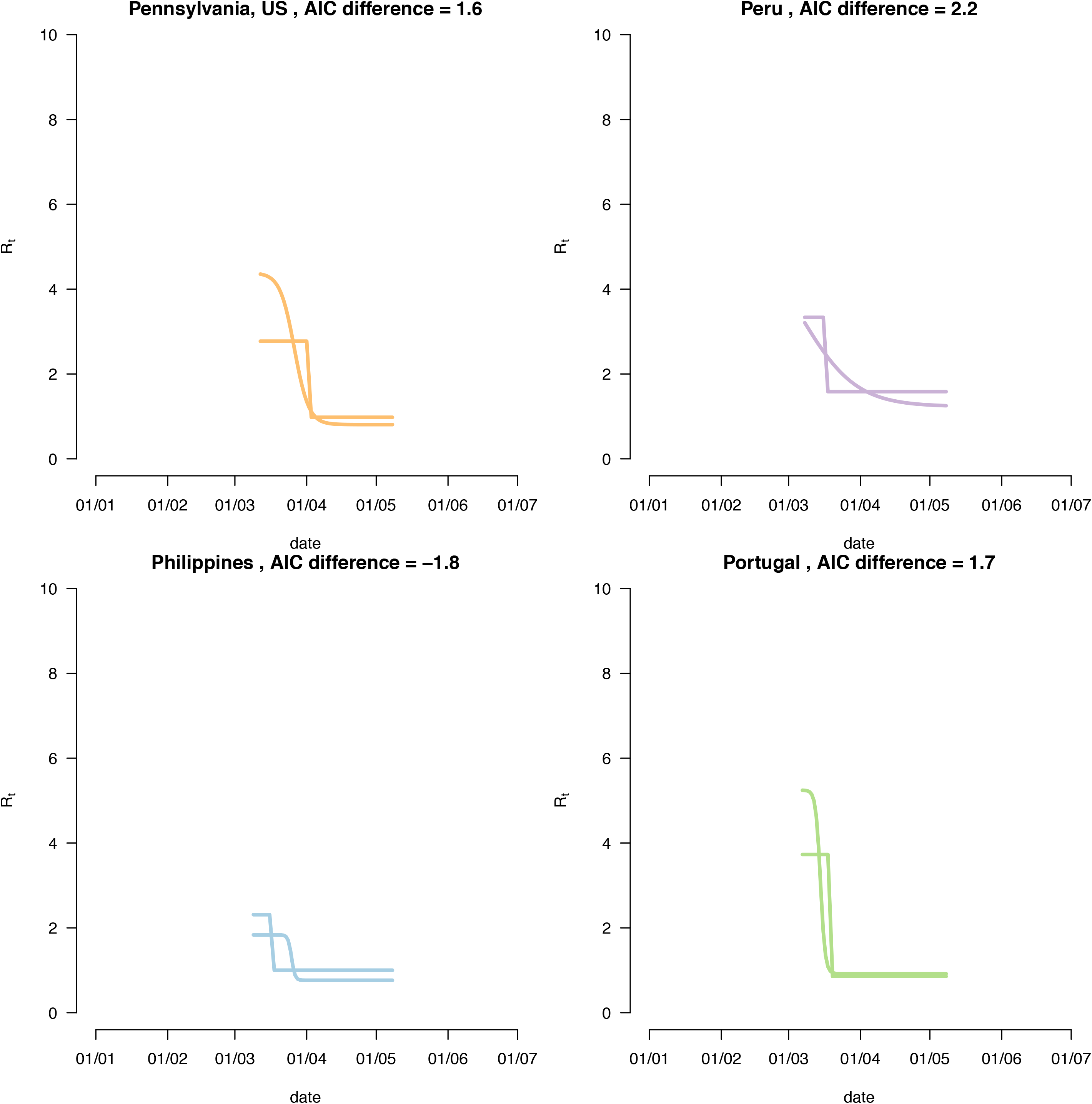
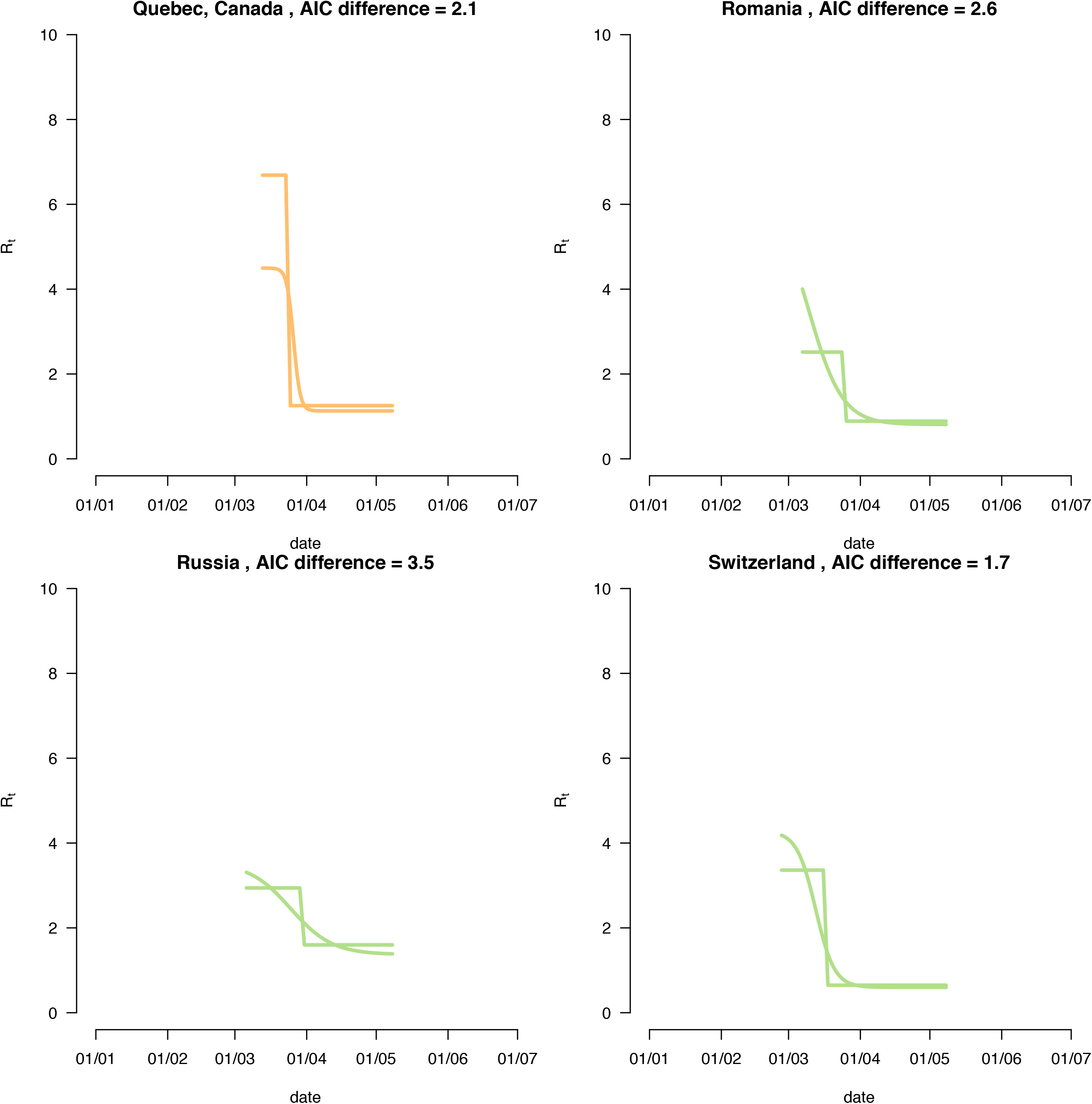
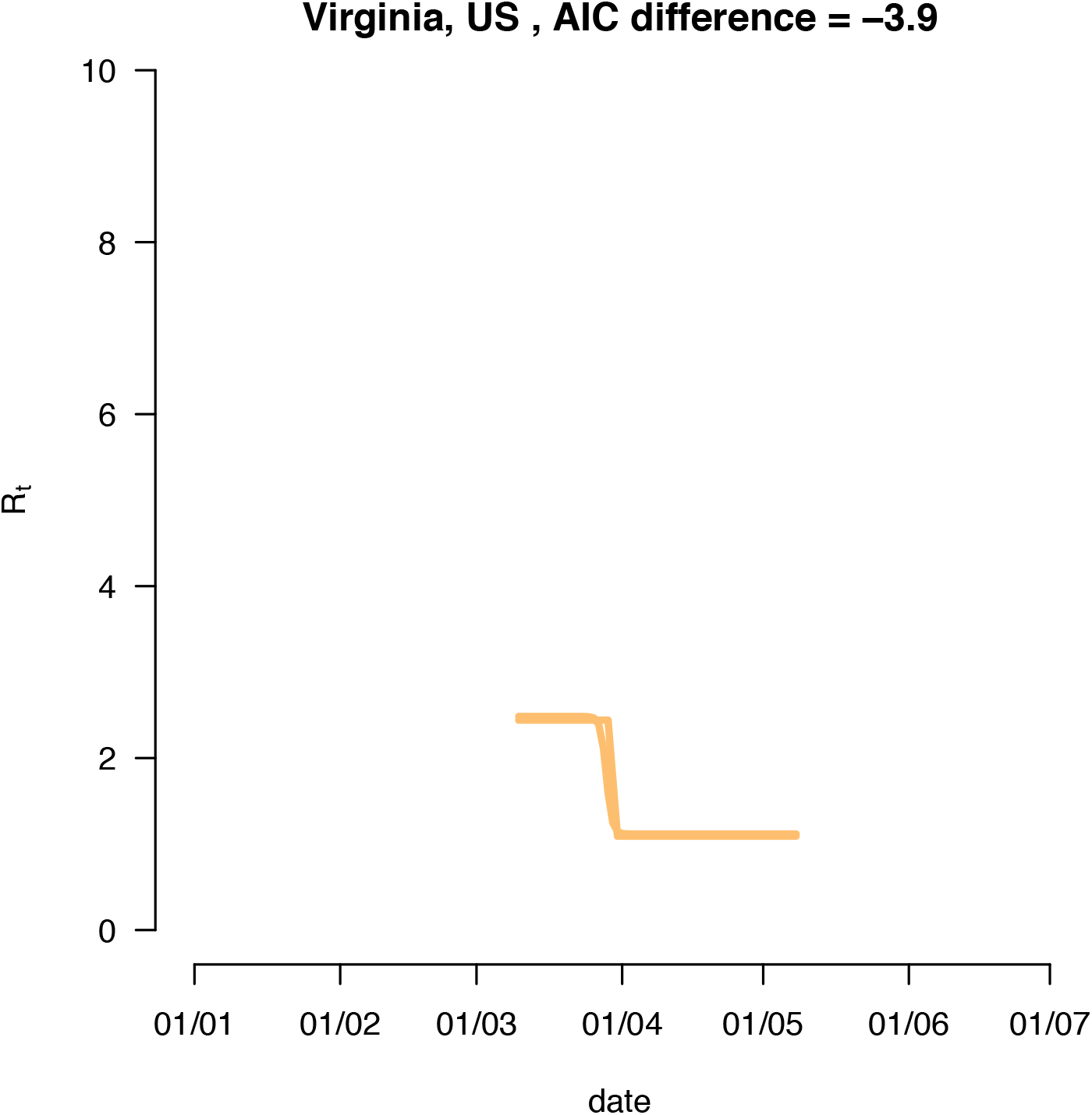
The transmissibility as a function of time for the sharp and the smooth decline in transmissibility, in the 29 regions where the smooth function did not fit the data better.

**Supplementary Figure 3:**
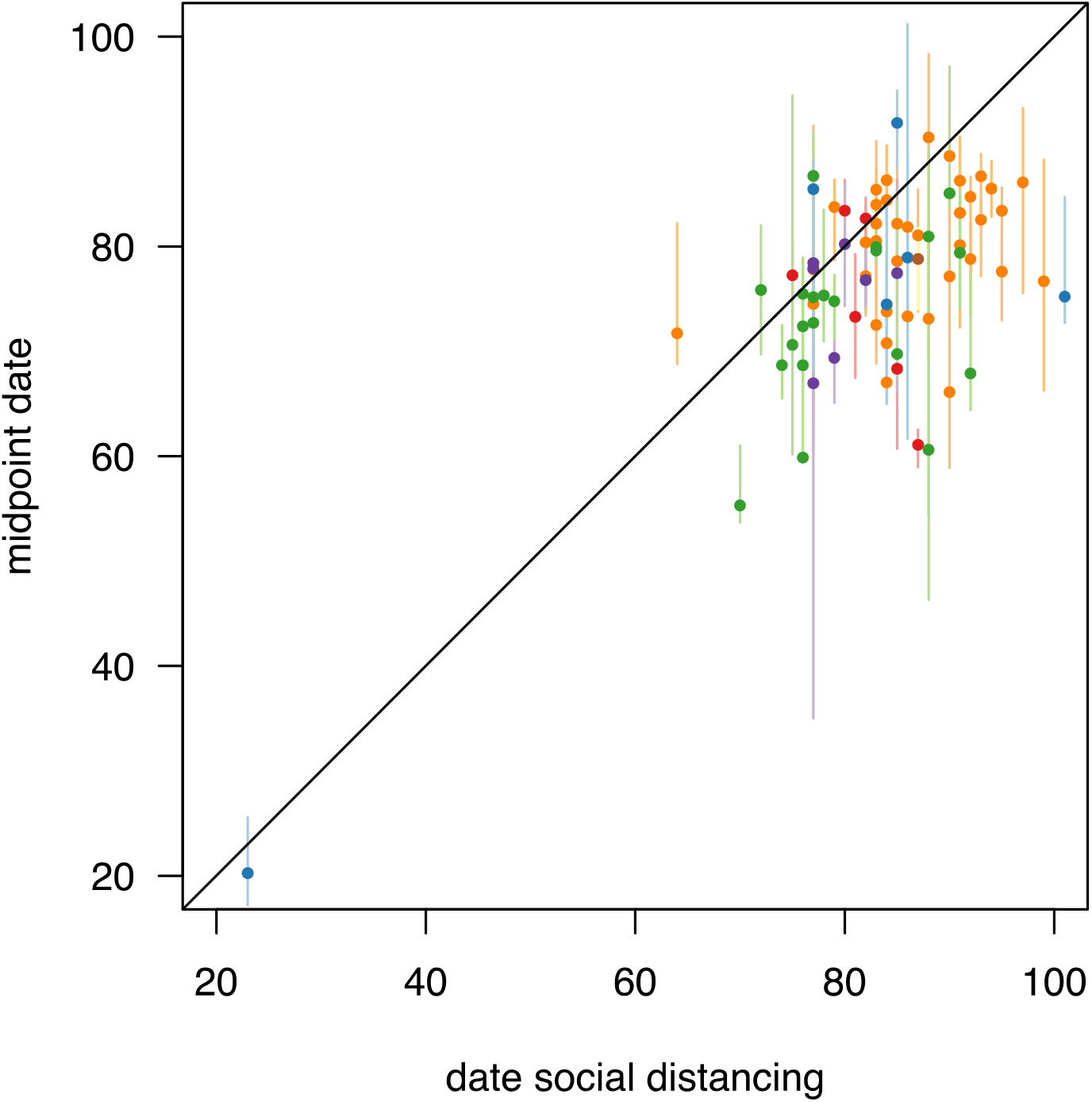
The correlation between the date of the lockdown and the inferred mid-point date of the smooth reduction in transmissibility. Date is in units of days in year 2020.

**Supplementary Figure 4:**
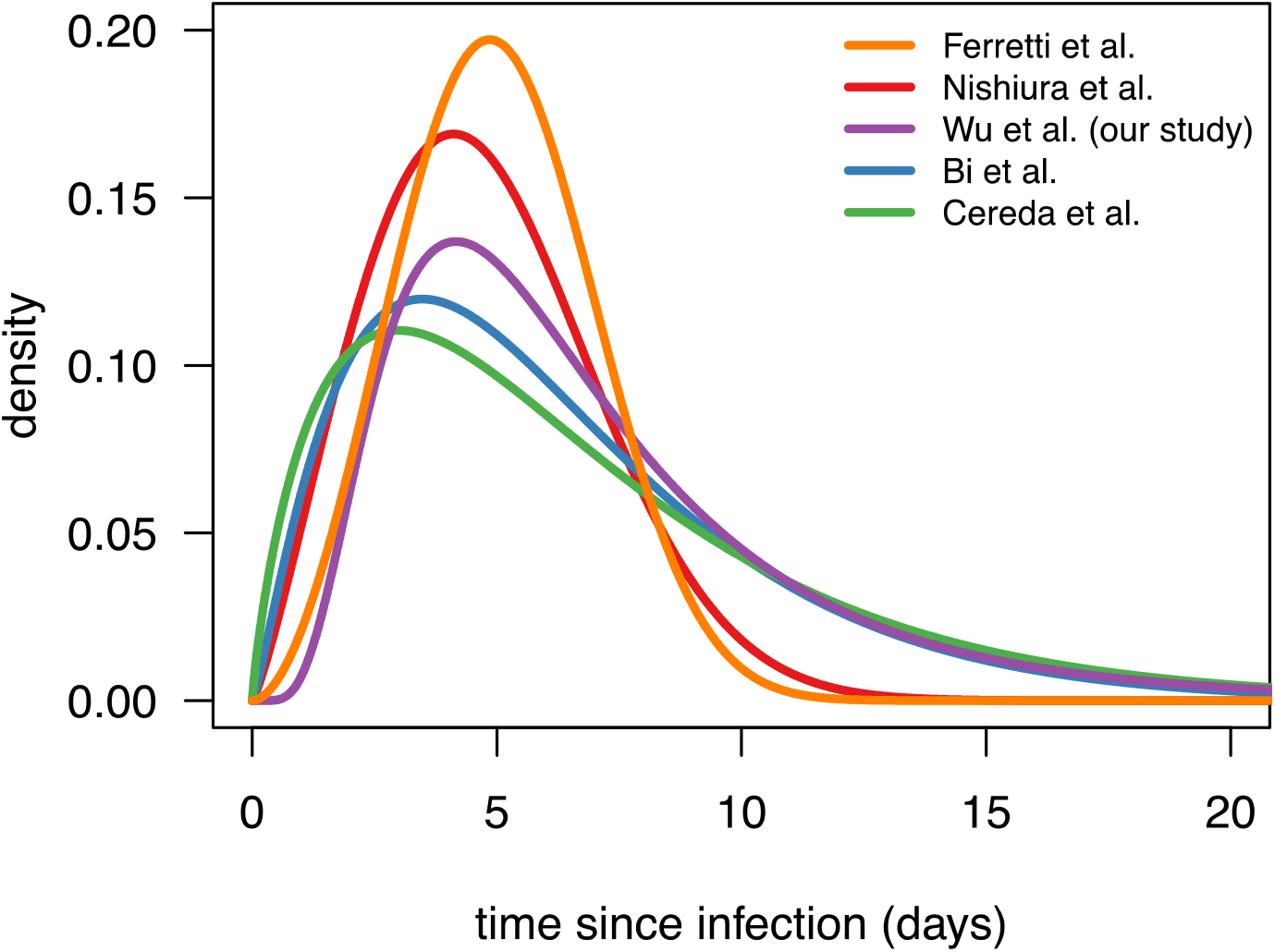
the inferred distributions of the serial inteval or generation time (Ferretti et al.) in five studies.

**Supplementary Figure 5:**
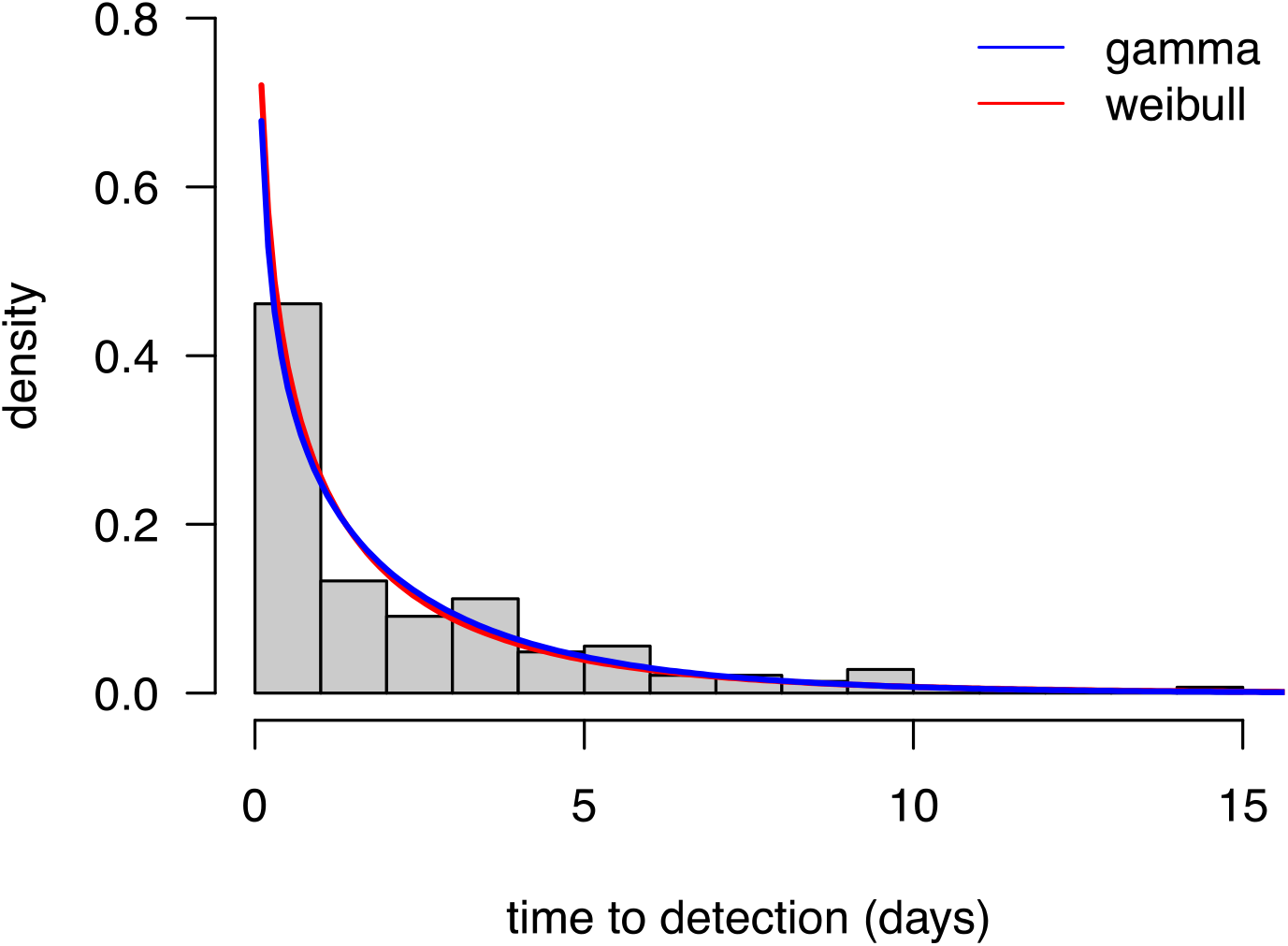
the distribution of the time from symptom onset to detection. The bars are the data on 150 cases, the curves are the fit of two distributions that fit the data equally well.

**Supplementary Figure 6:**
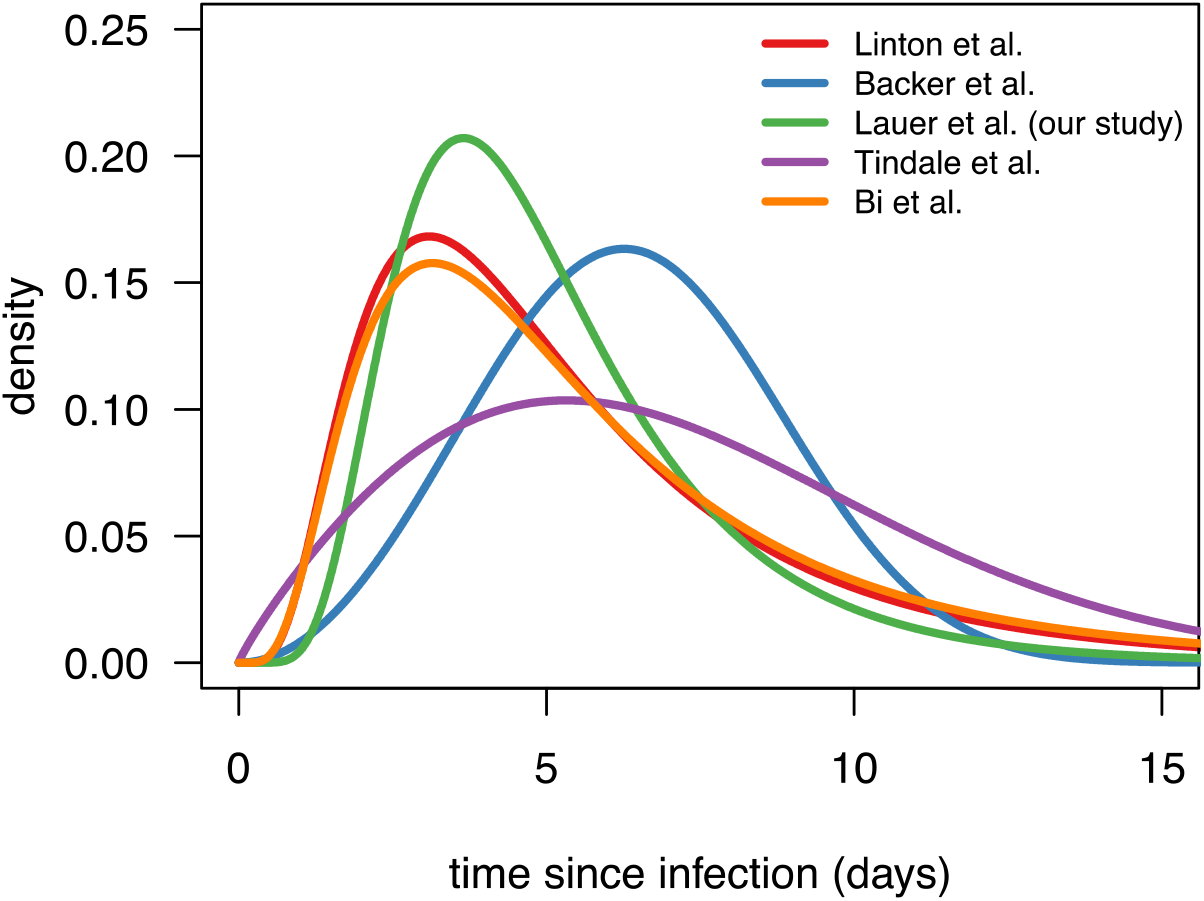
the inferred distributions of the time from infection to symptom onset in five studies. We use the relation in Lauer et al. 2020 (8).

Figure available at: https://github.com/FrancoisBlanquart/covid_model/blob/master/figureS7_merged.pdf]

Supplementary Figure 7: the daily cases (green) and deaths (blue) for all 79 regions examined, with the fit of three models (Model 0 based on deaths; model 1 based on cases and deaths and sharp reduction in transmissibility; model 2 based on cases and deaths and smooth reduction in transmissibility). The number of infected predicted by the model is shown in red. The insets show the inferred relative transmissibility and the inferred probability of detec ton over time. [figure available online].

**Supplementary Figure 8:**
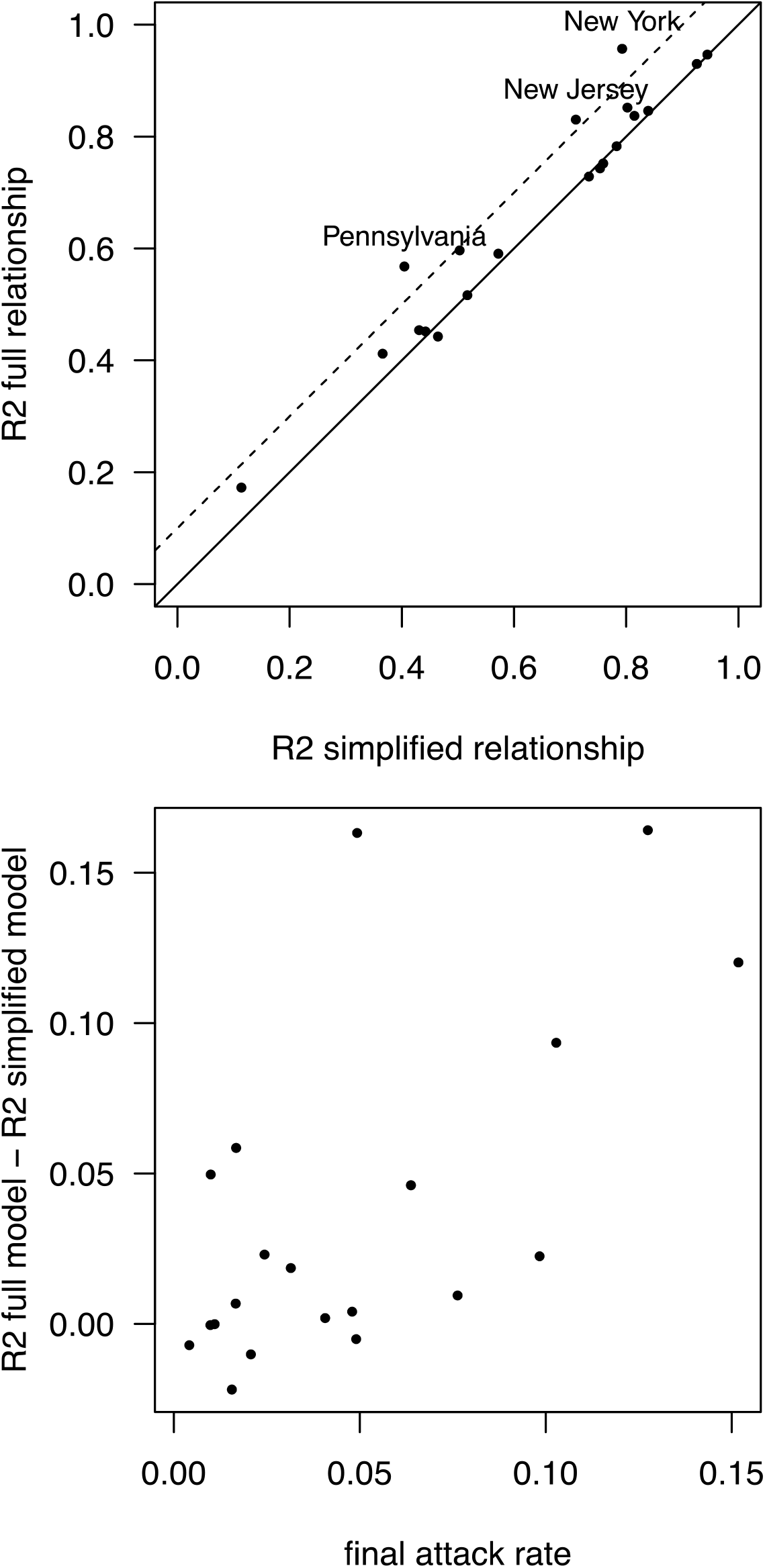
Top panel, comparison of the coefficient of determination of the general test model (equation (16)) with that of the simplified model (equation (17a)). The dashed line shows when the general model gives a coefficient of determination 10% better than the simplified model. Bottom panel, the improvement in fit of the general model positively correlates with the final attack rate.

